# A systematic review on outbreaks of COVID-19 among children within households in the European region

**DOI:** 10.1101/2022.10.17.22281168

**Authors:** Constantine I. Vardavas, Katerina Nikitara, Katerina Aslanoglou, Apostolos Kamekis, Nithya Ramesh, Emmanouil Symvoulakis, Israel Agaku, Revati Phalkey, Jo Leonardi-Bee, Esteve Fernandez, Orla Condell, Favelle Lamb, Charlotte Deogan, Jonathan E. Suk

## Abstract

**Objectives:** This systematic review aims to identify the secondary attack rates (SAR) to adults and other children when children are the index cases within household settings.

**Methods:** This literature review assessed European-based studies published in Medline and Embase between January 2020 and January 2022 that assessed the secondary transmission of severe acute respiratory syndrome coronavirus 2 (SARS-CoV-2) within household settings. The inclusion criteria were based on the PEO framework (P-Population, E-Exposure, O-Outcome) for systematic reviews. Thus, the study population was restricted to humans within the household setting in Europe (population), in contact with pediatric index cases 1–17 years old (exposure) that led to the transmission of SARS-CoV-2 reported as either a SAR or the probability of onward infection (outcome).

**Results:** Of 1,819 studies originally identified, 25 met the inclusion criteria. Overall, the SAR ranged from 13% to 75% in 23 studies, while there was no evidence of secondary transmission from children to other household members in two studies. Evidence indicated that asymptomatic SARS-CoV-2 index cases also have a lower SAR than those with symptoms and that younger children may have a lower SAR than adolescents (>12 years old) within household settings.

**Conclusions:** SARS-CoV-2 secondary transmission from paediatric index cases ranged from 0% to 75%, within household settings between January 2020 and January 2022, with differences noted by age and by symptomatic/asymptomatic status of the index case. Given the anticipated endemic circulation of SARS-CoV-2, continued monitoring and assessment of household transmission is necessary.

## Introduction

At the time of this review, Epidemiological data on severe acute respiratory syndrome coronavirus 2 (SARS-CoV-2) indicate that children are less prone to get infected by COVID-19 and, when infected, the clinical characteristics are less severe than those in adults (1). Virological studies of SARS-CoV-2, Middle East Respiratory Syndrome coronavirus (MERS-CoV) and SARS-CoV also suggest that children are less likely to develop serious illness following infection compared with adults (1). A significant area of respiratory research relates to the ability of infected children to infect others (2, 3). Previous research suggests that children are less frequently the index cases in both the household and school setting and are more likely to get infected by an adult. Higher rates of transmission have also been previously observed in older children (10–19 years old) in comparison to younger children (⍰<10 years old) (4).

To prevent the spread of COVID-19, social distancing policies within the first waves of the pandemic were instated, leading to the closure of educational settings within some countries and the requirement that children remain within households. In order to better understand the role of children in the transmission of SARS-CoV-2 outside the school setting, it is important to understand how SARS-CoV-2 was transferred within households during the COVID-19 pandemic. This would then be able to build the evidence for public health emergency preparedness actions for future pandemics in Europe. (5).

The aim of this systematic review is to identify the secondary attack rates (SAR) of adults and other children when children are the index cases within households in Europe up to January 2022.

## Methods

### Search strategy and selection criteria

A systematic literature review was performed in January 2022 according to the Preferred Reporting Items for Systematic Reviews and Meta-Analysis (PRISMA) guidelines (6). Relevant peer-reviewed studies were identified through systematic electronic searches using the OVID Medline and EMBASE databases. The complete search strategy and search terms are available in Supplementary Table 1.

The following set of inclusion criteria, based on adapted versions of the PEO framework (P-Population, E-Exposure, O-Outcome) for systematic reviews (7), was used to identify relevant studies and determine their eligibility for inclusion and are:

- Population: Humans, of any age within a household setting. The household setting includes cohabiting individuals, including family members, close relatives, or housemates.
- Exposure: Index cases, aged 0-17 years, defined as the first individual with laboratory-confirmed SARS-CoV-2 to develop symptoms or test positive within a household setting. Studies or reports that solely address non-household transmission were excluded.
- Outcome: Transmission of SARS-CoV-2 reported as either secondary attack rate (SAR, probability of onward infection from an index case among a defined group of close contacts), or observed reproduction number (R, observed average number of secondary cases per index case).
- Geographical Context: Europe, European Union (EU), United Kingdom (UK) and European Economic Area (EEA) countries.
- Study designs: All study types were considered, including descriptive studies, outbreak-cluster investigation reports, and contact tracing investigations. Systematic reviews and non-systematic reviews were identified, and references were screened for eligible studies. Opinion pieces and commentaries were excluded.
- Timeframe: 1 January 2020 to 20 January 2022

### Data analysis, extraction, tabulation and quality appraisal

Studies identified from the searches were uploaded into a bibliographic database, and duplicates were removed. Initially, a pilot training screening process was used, where a random sample of 100 titles was screened for eligibility independently by two reviewers [KA, AK] to enable consistency in screening and identify areas for amendments in the inclusion criteria. A high measure of inter-rater agreement was achieved (percentage agreement>80%), and hence the remaining titles were equally distributed between the two reviewers and screened independently. Any disagreements were thoroughly discussed with a third reviewer [CV]. For the full-text screening, a similar process was followed. Ten randomly selected studies were independently screened for eligibility by two reviewers [KA, AK] for the level of agreement to be estimated (percentage agreement>90%). The remaining full texts were equally distributed and screened by the two reviewers [KA, AK]. A data extraction template was independently piloted by two reviewers on a random sample of five included studies to assess consistency in data extraction and identify where amendments need to be made to the template. The remaining studies were then data extracted independently by the two reviewers [KA, AK]. Extracted data included study characteristics (first author’s name, year of publication), geographical context (country/area), methodology/study type, timeframe, setting (where the measures were implemented), COVID-19 diagnosis, contact tracing, SARS-CoV-2 strain, the non-pharmaceutical interventions (NPIs) implemented, follow up process, population characteristics (age, gender, geographical location), and objective and quantitative results with regards to transmission between children and/or transmission from children to adults, including Secondary Attack Rate (SAR) and Odds Ratios (OR).

The Joanna Briggs Institute (JBI) standardised critical appraisal tools were used for cohort studies, cross-sectional studies, case reports, and case series (8).

A narrative synthesis approach was followed, while where patterns in the data were identified through tabulation of results, an inductive approach (where concepts were derived from the data) was taken to translate the data to identify areas of commonality between studies. Where results are presented graphically, standard errors were either entered as reported or imputed based on the medians of the reported standard errors.

### Patient and public involvement

This study was performed under contract for the European Center for Disease Prevention and Control (ECDC). Patients or the public were not involved in the design, or conduct, or reporting, or dissemination plans of our research.

## Results

A total number of 1,819 studies were identified according to the specified selection criteria from the two databases. After removing duplicates, 1,788 were screened by title/abstract of which, 58 were assessed via full text. Of these 58 studies, 33 were excluded due to limited data, limited outcomes of interest, non-eligible geographical area, and irrelevant study type (reviews, conference abstracts, opinion papers). Hence, 25 studies were eventually considered in our analysis. The flowchart of study selection is presented below in **Figure 1**.

**Figure 1.**
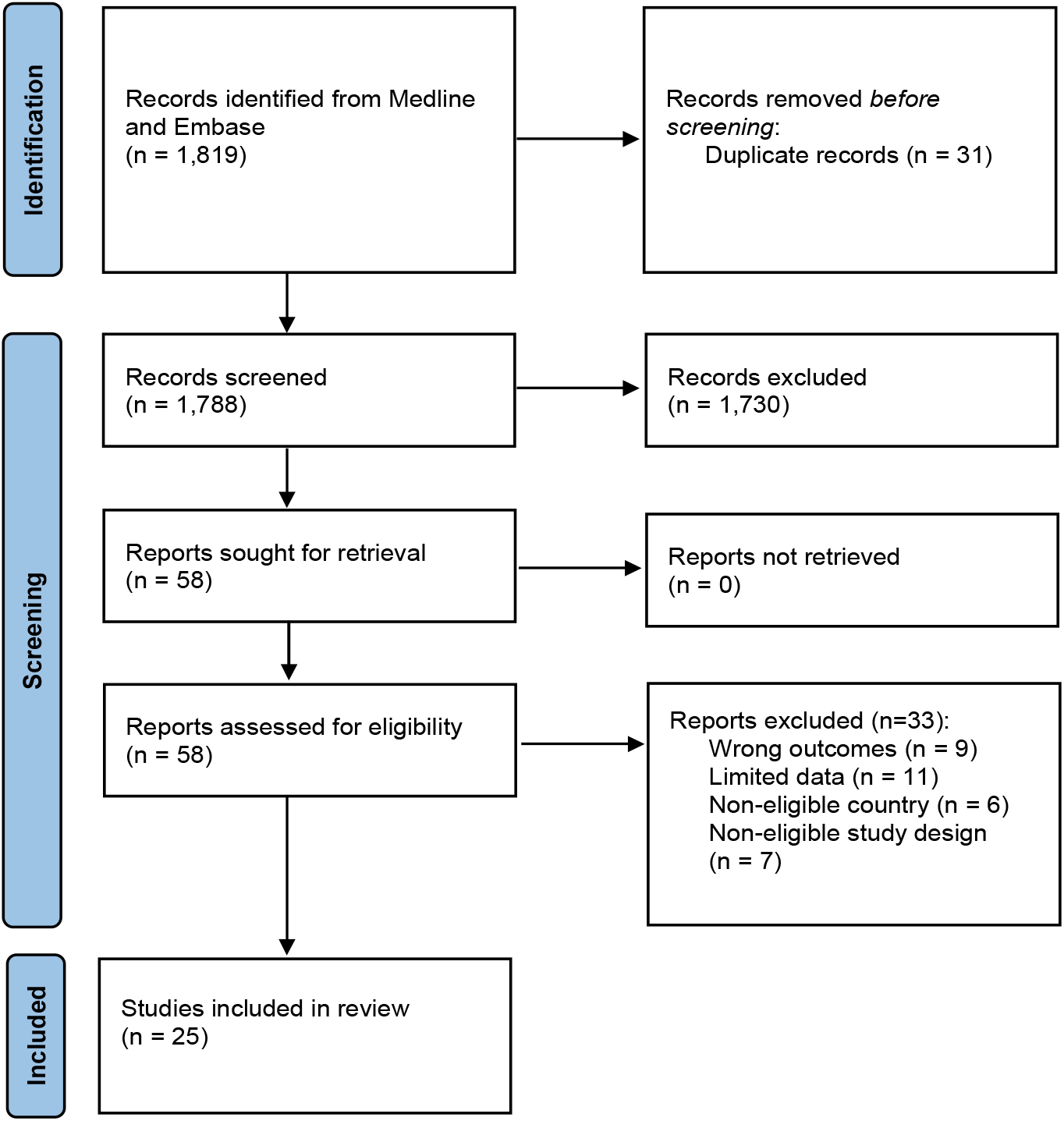
Flowchart of study selection

Of the 25 studies, 16 were cohort studies (9-24), five were cross-sectional (25-28), two were case reports (29, 30), one was a case series (31) and one was a case-control (32). Real-time polymerase chain reaction (RT-PCR) was used in 17 studies to diagnose COVID-19. In seven studies, serology tests were performed in addition to or instead of RT-PCR (10, 14, 22, 23, 26, 29, 32, 33), while in the remaining study, Nucleic Acid Amplification Test (NAAT) was reported as the single diagnostic method for SARS-CoV-2 detection (25). Regarding NPIs, case isolation and quarantine of contacts were the most frequently reported measures and implemented in parallel with the contact tracing investigations for mitigating/suppressing SARS-CoV-2 transmission during the pandemic.. These features along with the geographic area of the study are summarised in **Supplementary Table 2**.

### Child to adult/child SARS-CoV-2 transmission in the household setting

Of the 25 studies included in the analysis, 18 provided adequate contact tracing data for estimating the SAR from paediatric index cases to adult and/or child household contacts (**Table 1**). Overall, SAR ranged from 13% to 75% in 23 studies, while in two studies there was no evidence of secondary transmission from children to other household members (18, 20). The highest SARs (67% and 75%) were found in studies examining single-family clusters with symptomatic paediatric index cases (29, 30). In studies where an age stratification was performed (n=7 studies), higher SARs were mostly noted from adolescents (>12 years old) (19, 22, 27, 28, 32) compared to younger ages, except for two studies, the results of which indicated higher SAR from children 0 to 11 years old (9, 24).

**Table 1.**
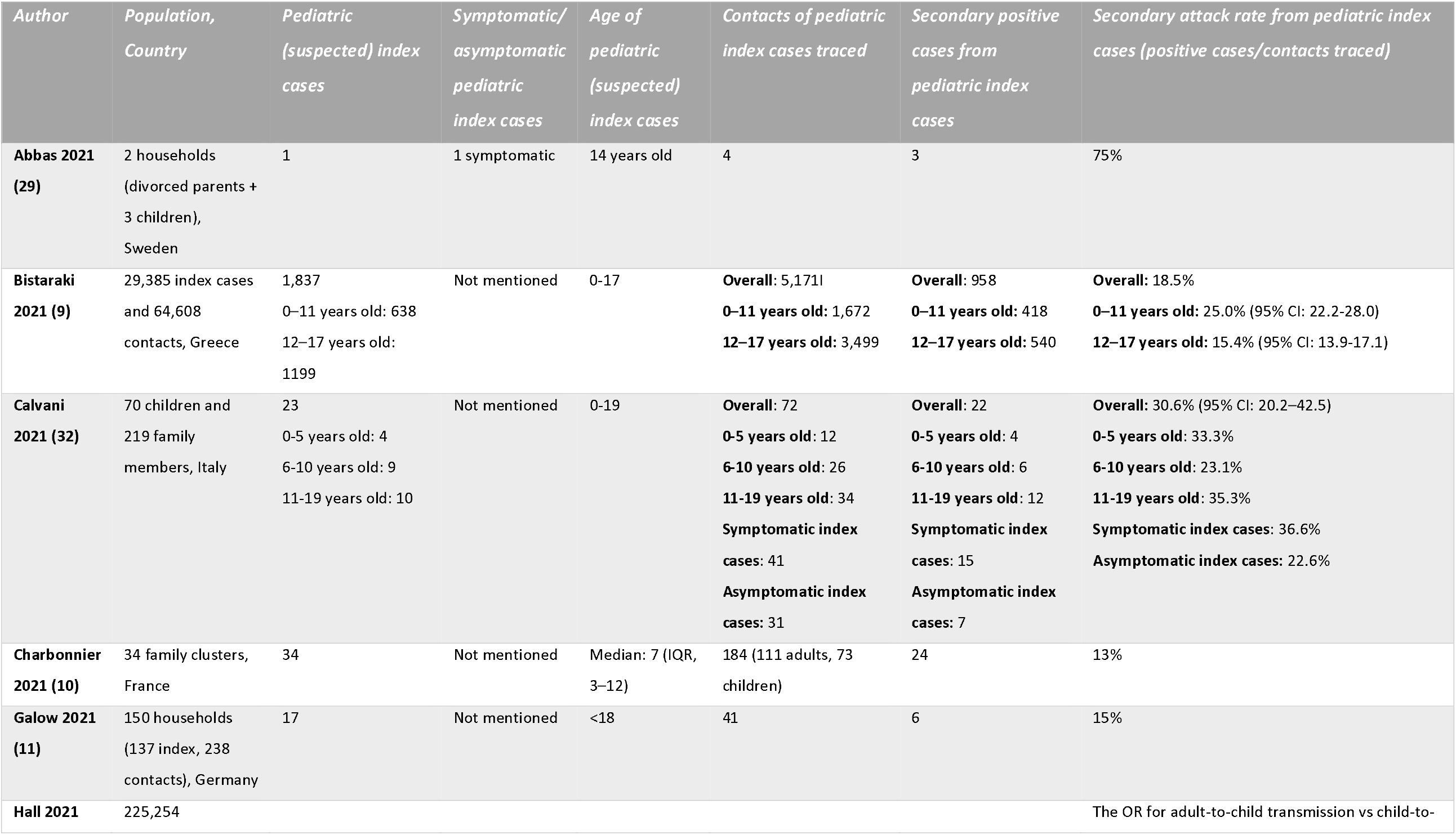
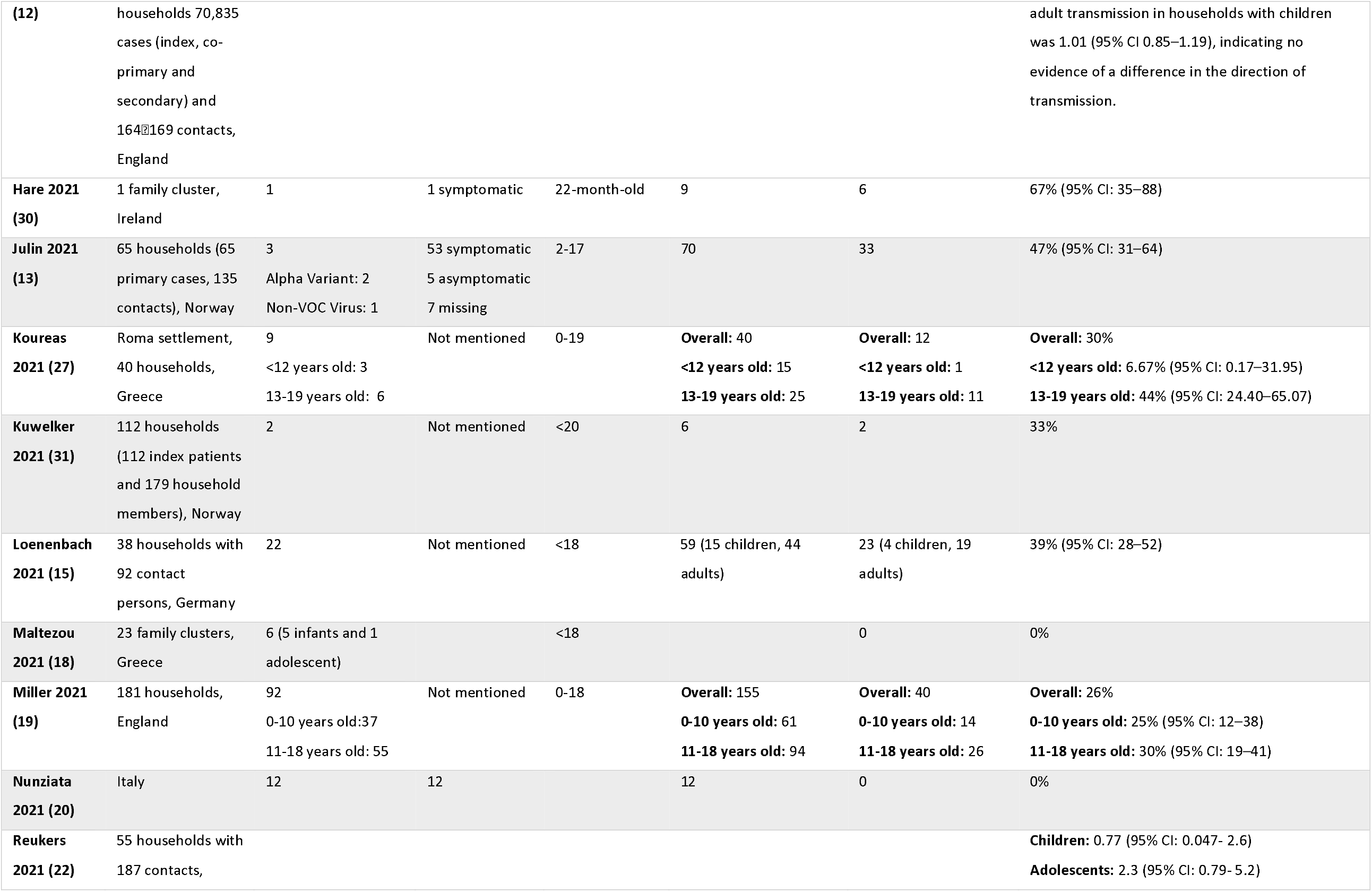
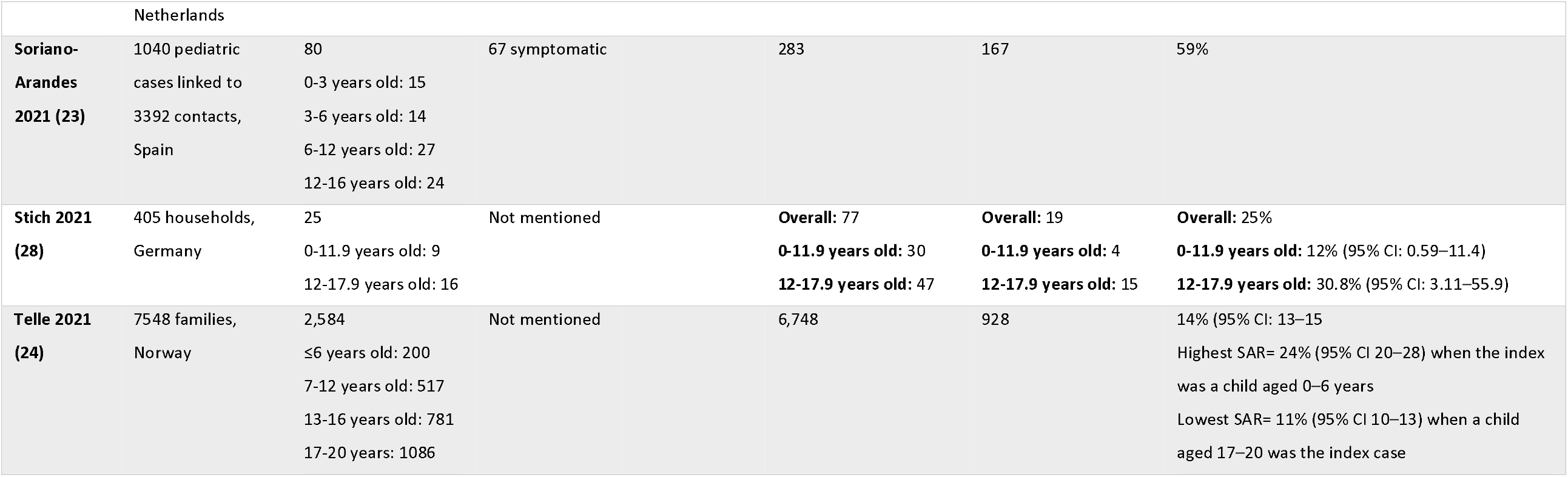
Assessment of secondary attack rates from pediatric index cases.

The highest SAR was found in the study of Abbas et al. (2021) (29), tracing a family cluster residing in Sweden. Among the four family contacts of the paediatric index case, three secondary infections occurred, with the SAR being estimated at 75%. A high SAR was found in a family cluster from Ireland, as described by Hare et al. (2021) (30), where SARS-CoV-2 was transmitted from one symptomatic paediatric index case to six of nine household contacts, leading to a SAR of 67% (95% CI: 35-88). A slightly decreased SAR of 59.0% was presented by Soriano-Arandes et al. (2021) (23) in a study performed in Catalonia, Spain with 80 paediatric index cases, 67 of which were symptomatic and transmitted SARS-CoV-2 to 167 out of 283 household contacts.

A more meticulous approach for SAR based on age stratification was provided by seven studies. Calvani et al. (2021) (32) performed a case-control study to investigate 70 pediatric cases, 28 of which were reported as index cases in their households. The overall SAR from children to household members was estimated at 30.6% (95% CI: 20.2-42.5), while the highest SAR was found in the ages of 0-5 (33.3%) and 11-19 years old (35.3%) compared to index cases aged 6-10 years old (23.1%). The study also showed that the 80% of symptomatic paediatric index cases spread the virus to their family members compared to 26.7% for asymptomatic paediatric index cases (SAR: 36.6% versus 22.6%, respectively).

The predicted SAR in 77 exposed household members was lower when the index case-patient was <12 years of age (SAR=12.0% (95% CI: 0.59-11.4)) and higher with an index case-patient 12-17.9 years of age (SAR=30.8% (95% CI: 3.11-55.9)). Moreover, transmission rate was estimated at 0.77 (95% CI: 0.047-2.6) per infectious period for children, and at 2.3 (95% CI: 0.79-5.2) for adolescents by Reukers et al. (2021) in households with a median of 4 (3-9) persons (22). Compared with the previous findings, Bistaraki et al. (2021) (9), who studied 1,837 pediatric index cases, estimated a higher SAR from children at the age of 0-11 years (SAR=25% (95% CI: 22.2-28)) compared to adolescents (SAR=15.4% (95% CI: 13.9-17.1)). Likewise, in the study by Telle et al. (2021) (24), SAR was 24% (95% CI 20–28) when the index was a child aged 0–6 years and declined with increasing age of the index child, with the lowest SAR (11%, 95% CI 10–13) found when a child aged 17–20 was the index case.

The lowest SARs, ranging from 13% to 15% were detected by Charbonnier et al. (2021)^9^ and Galow et al. (2021) (11), while Maltezou et al. (2020) (17) and Nunziata et al. (2021) (20) found no secondary SARS-CoV transmission from paediatric index cases. Similarly, in the study published by Maltezou et al. (2021) (18) none of the six paediatric index cases from 23 family clusters transmitted SARS-CoV-2 to any of the household contacts, while children were more likely to have an asymptomatic SARS-CoV-2 infection compared to adults (40% vs 10.5%; P⍰= ⍰.021) and were significantly more likely to have a low viral load. Finally, in the study of Hall et al. (2021) (12) the (OR) for adult-to-child transmission vs child-to-adult transmission in households with children was 1.01 (95% CI 0.85–1.19), indicating no evidence of a difference in the direction of transmission. **Figure 2** provides a graphical overview of the SAR when children are the index cases for studies which had available data reported as percentages and provided information on the standard errors.

**Figure 2.**
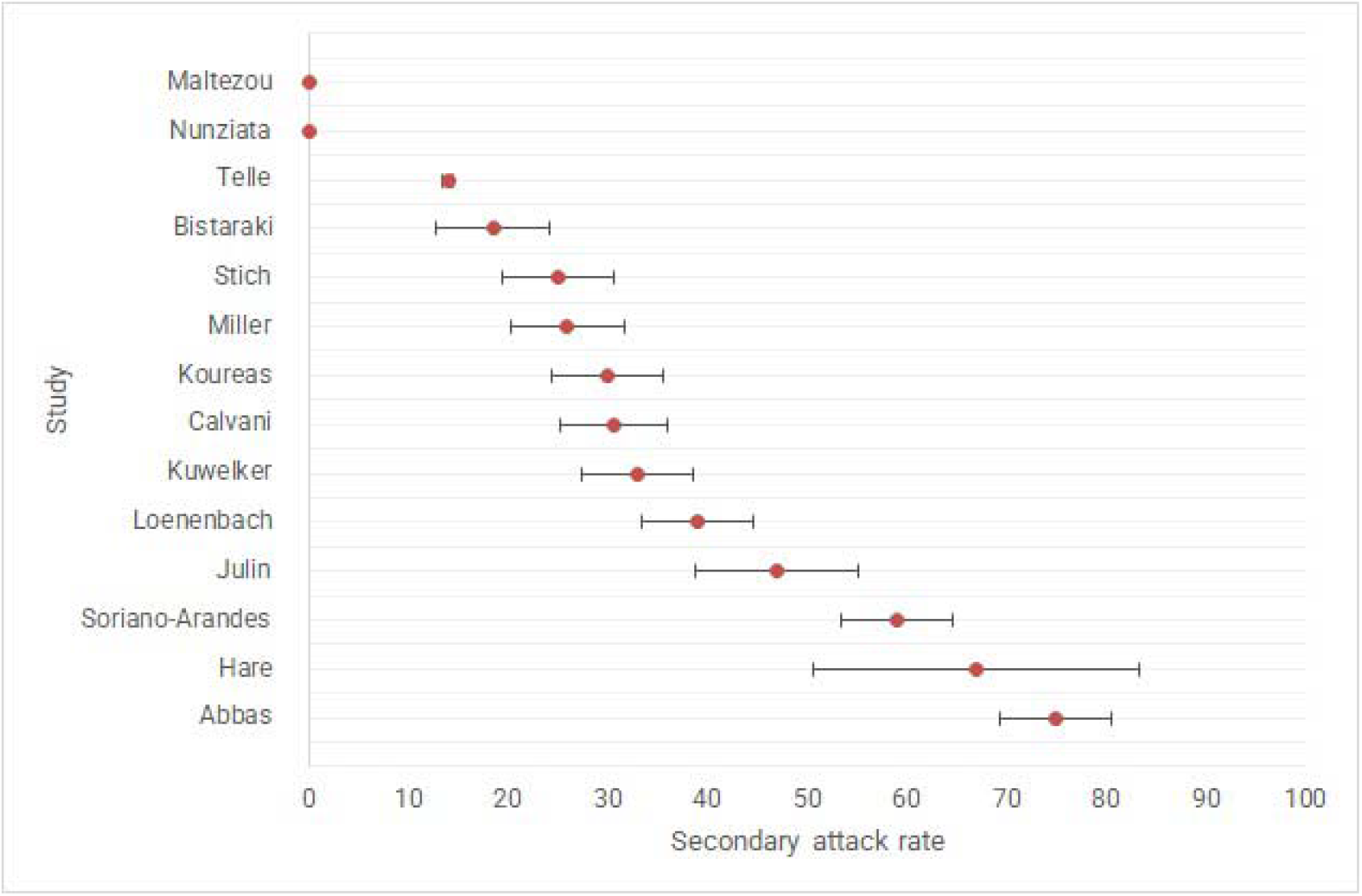
Graphical overview of the studies reporting a secondary attack rate (SAR) in European households when children are the index case. Footnote: Confidence intervals are either reported as is or imputed from the reported standard errors.

### Information on the frequency of pediatric index cases in household settings

The evidence on the frequency of paediatric index cases in household settings is detailed in Table 2. The highest proportion of paediatric index cases was detected in the studies conducted by Chudasama et al. (2021) (33), Loenenbach et al. (2021) (15), Miller et al. (2021) (19) and Maltezoy et al. (2020) (18), ranging from 47% to 59%. Chudasama et al. (2021) (33) found 13,215 paediatric index cases in a total number of 22,538 households, of whom 5,476 were at the age of 5-11 years and 7,739 at the age of 12-15 years. The authors reported that the proportion of household clusters where a child (aged 5–15 years) was identified as the index case remained similar over the summer of 2021. Similarly, Loenenbach et al. (2021) (15) conducted contact tracing among 38 households in which, 22 households had children who developed symptoms of COVID-19 and were assumed as the suspected index cases. Maltezou et al. (2020) (17) also investigated 187 index cases among 133 family clusters of which 62 were paediatric cases. Of these 62 children, 51 had no other family member with a SARS-CoV-2 infection, one child had an unknown family history and 10 children had at least one family member with a SARS-CoV-2 infection.

**Table 2.**
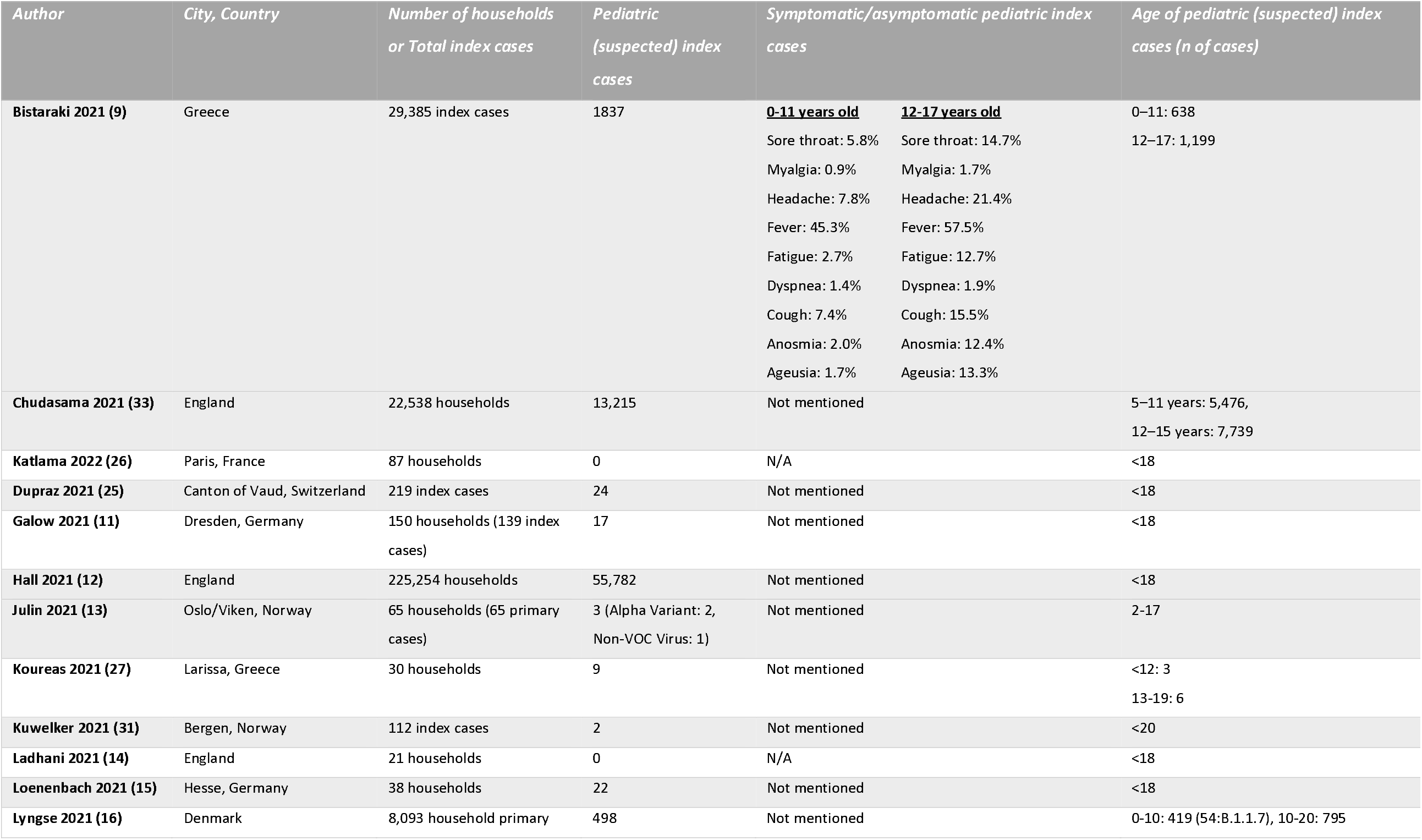
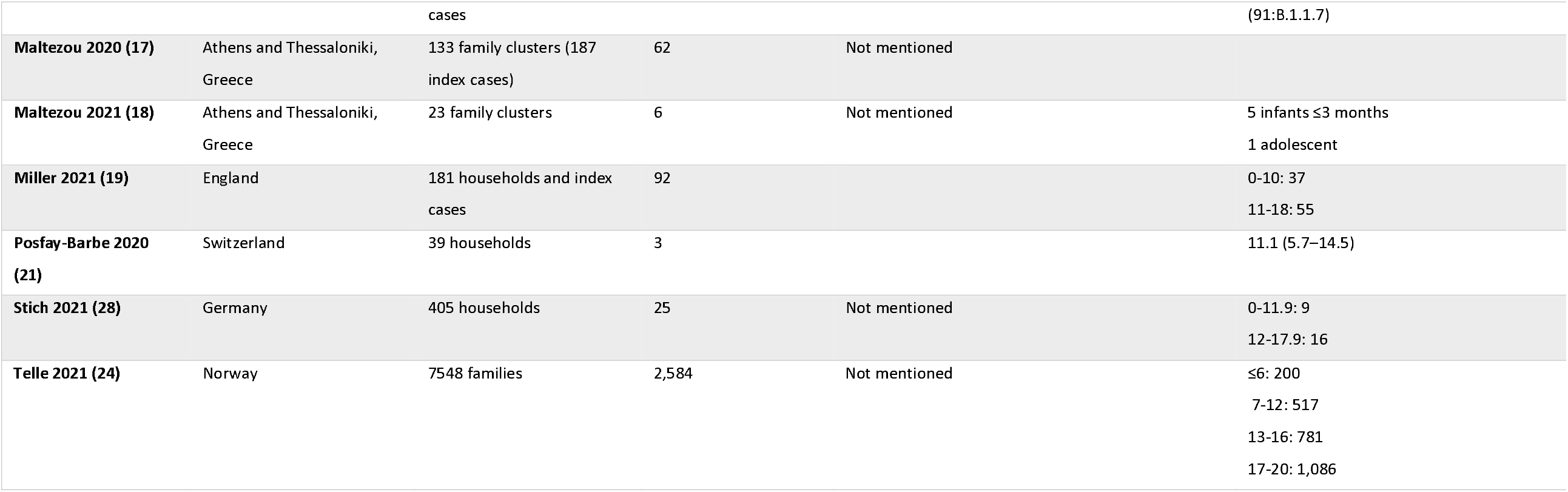
Study characteristics and results for studies with children and adults as index cases.

A lower proportion of paediatric index cases was found in the study of Telle et al. (2021) (24), who included 7,548 index cases, among which 4,964 were parents (66%) and 2,584 children (34%). From those 2,584 children, the number of index cases increased steeply with age, from 200 (8% of child index cases) among the youngest (aged 0–6) to 1,086 (42% of child index cases) among the oldest (aged 17–20). An increasing number of index cases with age was also noted by Koureas et al. (2021) (27), who reported three index cases younger than 12 years old and six at the age of 13-19 years. An estimated percentage of approximately 25% paediatric index cases was found in the samples of Maltezou et al. (2021) (17) and Hall et al. (2021) (12) in 23 family clusters and 225,254 households, respectively. In the remaining eight studies children were less frequently reported as index cases, whereas in two studies no paediatric index cases were found (14, 26). Lyngse et al. (2021) (16) investigated SARS-CoV-2 variant B.1.1.7 (Alpha variant) and other lineages and found among 8,093 primary household cases, 1,293 were children and adolescents of which 145 were diagnosed with the Alpha variant. The same variant was also detected in two out of three paediatric primary cases from 65 households in the study of Julin et al. (2021) (13). Finally, among the lowest proportions of paediatric index cases were found by Stich et al. (2021) (28) (25/405 households) and by Kuwelker et al. (2021) (31) (2/112 index cases). In the study by Stich et al. (28) adolescents were more frequently reported as index cases compared to younger children, like in all other studies where an age stratification was performed.

## Discussion

This systematic review provides an assessment of the peer-reviewed literature pertaining to SARS-CoV-2 transmission by children, within the household setting in the European context. Evidence suggests that transmission of SARS-COV-2 is higher in household settings than in other community microenvironments, including schools. This finding is potentially attributable to the individual, behavioural and contextual factors of households (34). The literature appraised in this review provides sufficient evidence that children less than 12 years old are less frequently reported as index cases in their households compared to adults, with adolescents showing a higher frequency of being the index case among the included studies than children under 11 years of age. This finding is corroborated by research published before the cut-off date of this review, estimating the overall weighted prevalence of parents being the index case of COVID-19 at 54% (35).

Regarding the transmissibility of SARS-CoV-2 from children to other household members, the SARs detected in the studies of this review ranged between 13% and 75%, while two studies showed no secondary transmission from child index cases (18, 20). Our results are similar to a recent meta-analysis of 18 studies with a more global scope, which estimated the SAR of child index cases at 20% (95% CI: 15–26, I2 = 100%), indicating that child index cases were significantly associated with a lower possibility to transmit SARS-CoV-2 to their family members (RR = 0.64, 95% CI: 0.50–0.81, I2 = 96%) compared with the adult index cases (36). A previous review also presented pooled estimates of an overall SAR from children at 10% (95% CI 3-25), and identified a lower child-to-child transmission rate at 5.7%, whereas the child-to-adult transmission rate was 26.4% (37). With regard to the differences noted between the estimated SARs of these reviews, it should be taken into account that emerging SARS-CoV-2 variants of concern have increased transmissibility, population vaccination rates have increased (38), and NPIs can be implemented differently across countries, which may explain differences in the identified SARs by setting. It is further interesting to note that children under 12 have lower vaccination rates compared to adults, on average, according to the EU vaccine tracker - a fact which may further influence household transmission (39).

There is evidence of differing transmission dynamics between younger vs older children, e.g. index cases under 11 years of age lead to lower SARs than older children. Moreover, although children appear to be at lower risk for symptomatic disease, symptomatic index cases had significantly higher SAR compared to asymptomatic index cases. Our results align with those of Chen et al. (2022), where symptomatic index cases were associated with a higher SAR than asymptomatic index cases (36).

Apart from the role of the child’s age in the household transmission of SARS-CoV-2, there are also environmental and behavioural factors which might facilitate or prevent secondary infections, including, but not limited to, the number of household members, the number of people per room, non-compliance with isolation requirements, sharing of index case’s bedroom, sharing of meals, as well as the level of adherence to facemask wearing (26, 40).

Finally, the current review identified a higher SAR within households when compared to the results of our previous review that assessed the transmission in educational settings, which noted limited cases of extensive secondary transmission in schools, especially when social distancing measures, facemasks and adequate ventilation were implemented (5).

### Strengths and limitations

There are limitations to this study that may impact the implications for decision-making. As we assessed peer-reviewed evidence published in two biomedical databases, it inherently reflects the status quo of the interim of the years (2020 - 2021) due to the lag time between study implementation, peer review and publication. Moreover, we report on studies that represent child-to-child/adult transmission within the context of initial SARS-CoV-2 strains and are not directly applicable to newer variants, such as the SARS-CoV-2 Delta or Omicron variant. Although we restricted studies to only those that were located within the EU/UK/EEA region so as to enhance comparability, the household transmission may also be influenced by other factors such as background levels of community SARS-CoV-2 transmission, the transmission of SARS-CoV-2 in educational settings (5) and varying NPI policies. Another matter of inconsistency is the different definitions of primary and index cases used in the included studies, as well as the various methods used for the identification of index cases, the contact tracing process and the follow-up duration. Supporting educators and parents in the implementation of NPIs may be important as population-based studies have indicated that adults concerned about the impact of COVID-19 on their children’s education may be more likely to practice personal protective measures and social distancing (41).

## Conclusion

According to the findings of studies that have been published up until January 2022, which in principle represent evidence from the first two years of the pandemic, the role of children in COVID-19 transmission within the household setting in the European region was notable, but higher than SARs noted in educational settings. Moreover, there was an indication that younger children may have a lower SAR than adolescents within household settings. Moreover, symptomatic paediatric index cases had significantly higher SAR than asymptomatic index cases. However, there were insufficient data to examine how the transmissibility of paediatric index cases is affected by different SARS-CoV-2 variants as well as the effect of vaccination on the spread of SARS-CoV-2 within the household setting. Given the potential endemic circulation of SARS-CoV-2, continued monitoring and assessment of household transmission is necessary.

## OTHER FIELDS REQUESTED

### Strengths and limitations of this study

- This review followed a systematic search approach.
- The search represents peer-reviewed literature that included previous SARS-CoV-2 variants but does not cover the Delta or Omicron variants.
- Our results were restricted to the European region so as to enhance comparability for policymakers and hence may not be reflective of the status quo in other regions as household transmission may also be influenced by other factors such as background levels of community SARS-CoV-2 transmission, the transmission of SARS-CoV-2 in educational settings and varying NPI policies.

## Data Availability

Data sharing is not applicable as no datasets were generated and/or analysed for this study.

## Funding statement

This report was commissioned by the European Centre for Disease Prevention and Control (ECDC) to the PREP-EU Consortium, coordinated by Dr Constantine Vardavas (School of Medicine, University of Crete) under Framework contract ECDC/2019/001 Lot 1B.

## Competing interests statement

The authors declare that they have no known competing financial interests or personal relationships that could have appeared to influence the work reported in this paper.

## Data availability statement

Data sharing is not applicable as no datasets were generated and/or analysed for this study.

## Acknowledgements

We would like to thank Katerina Papathanasaki (UoC), Chrysa Chatzopoulou (UoC) and Konstantinos Skouloudakis (UoC) for their assistance in data archiving.

## Supplementary Material

**Supplementary Table 1:**
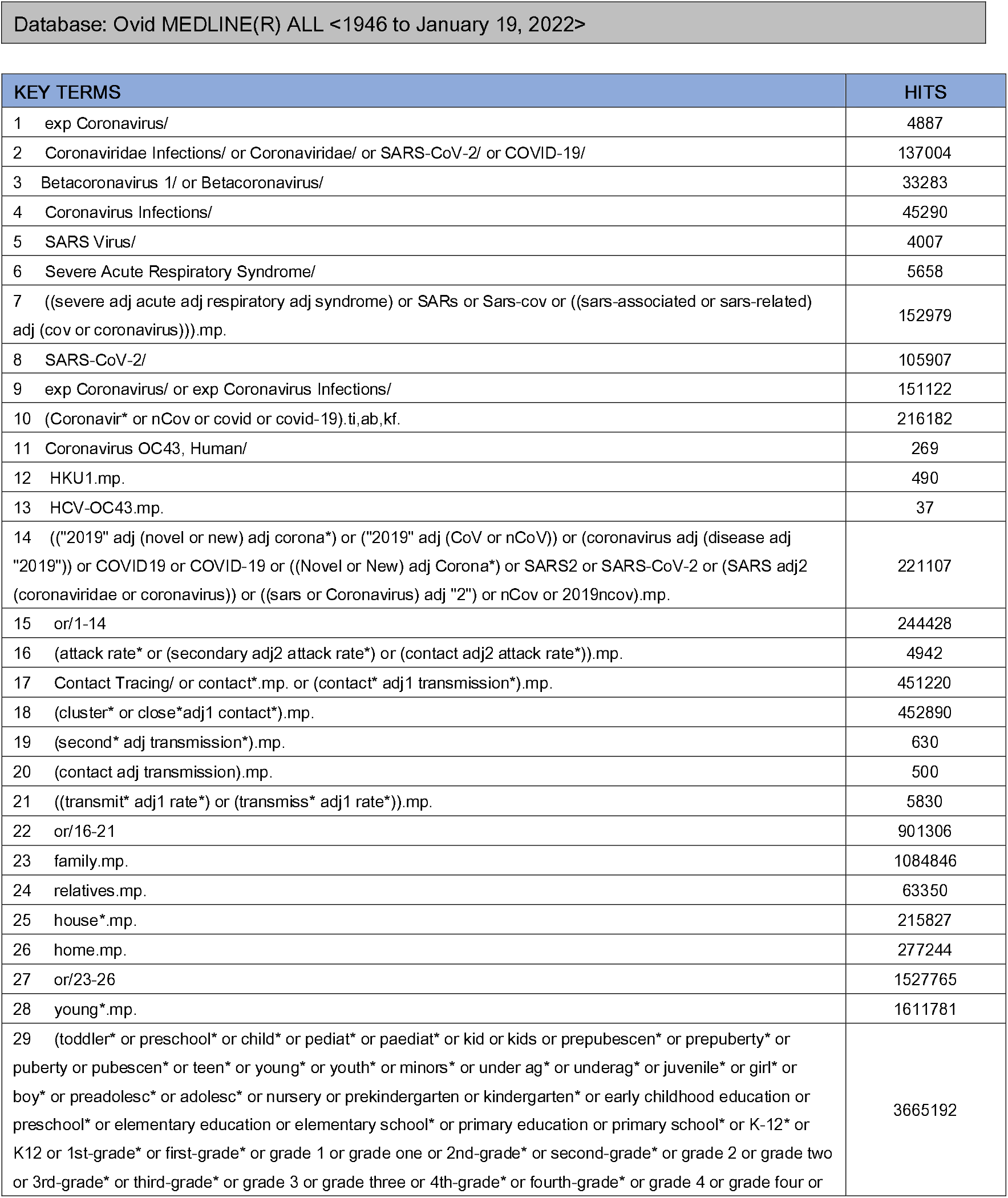
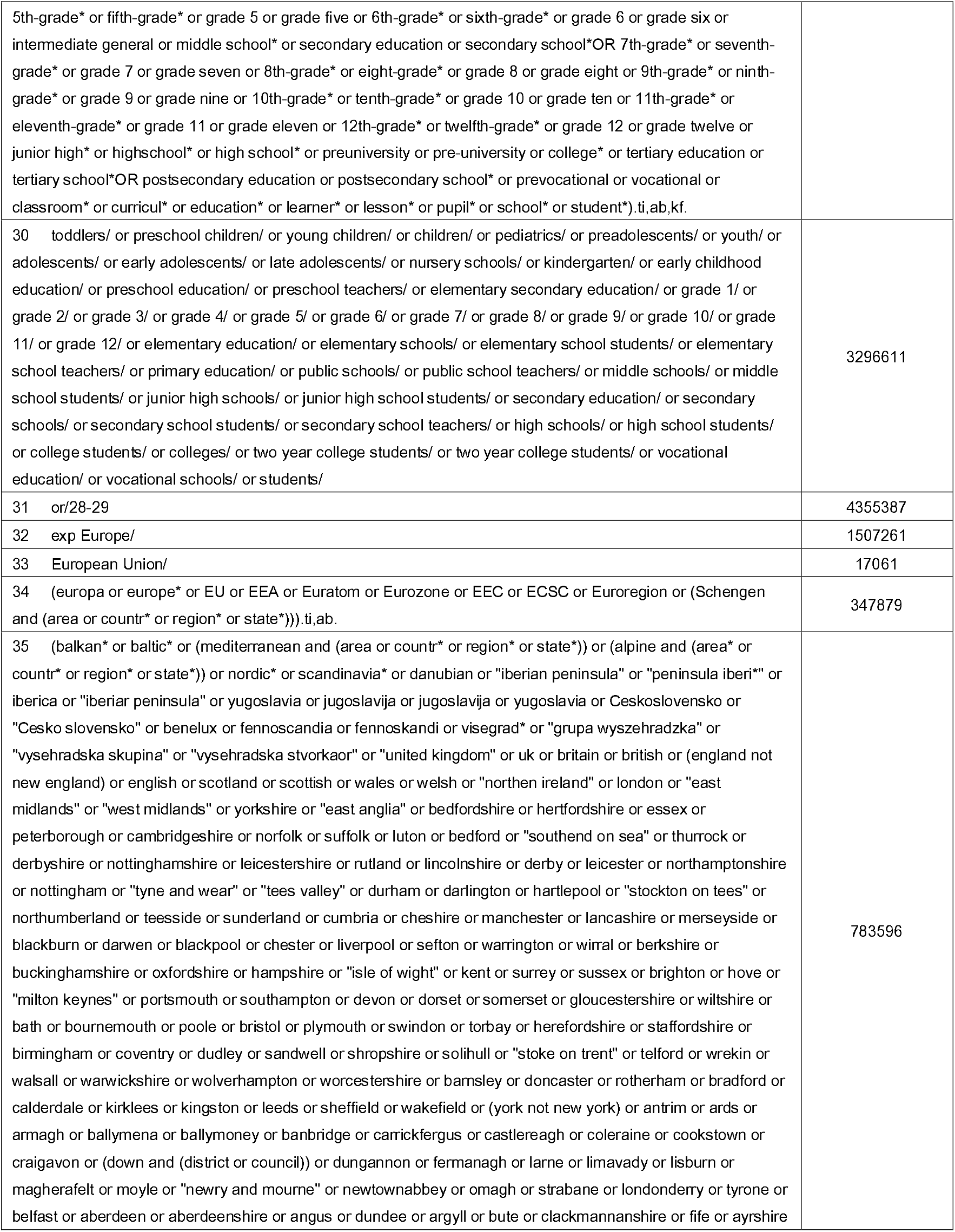
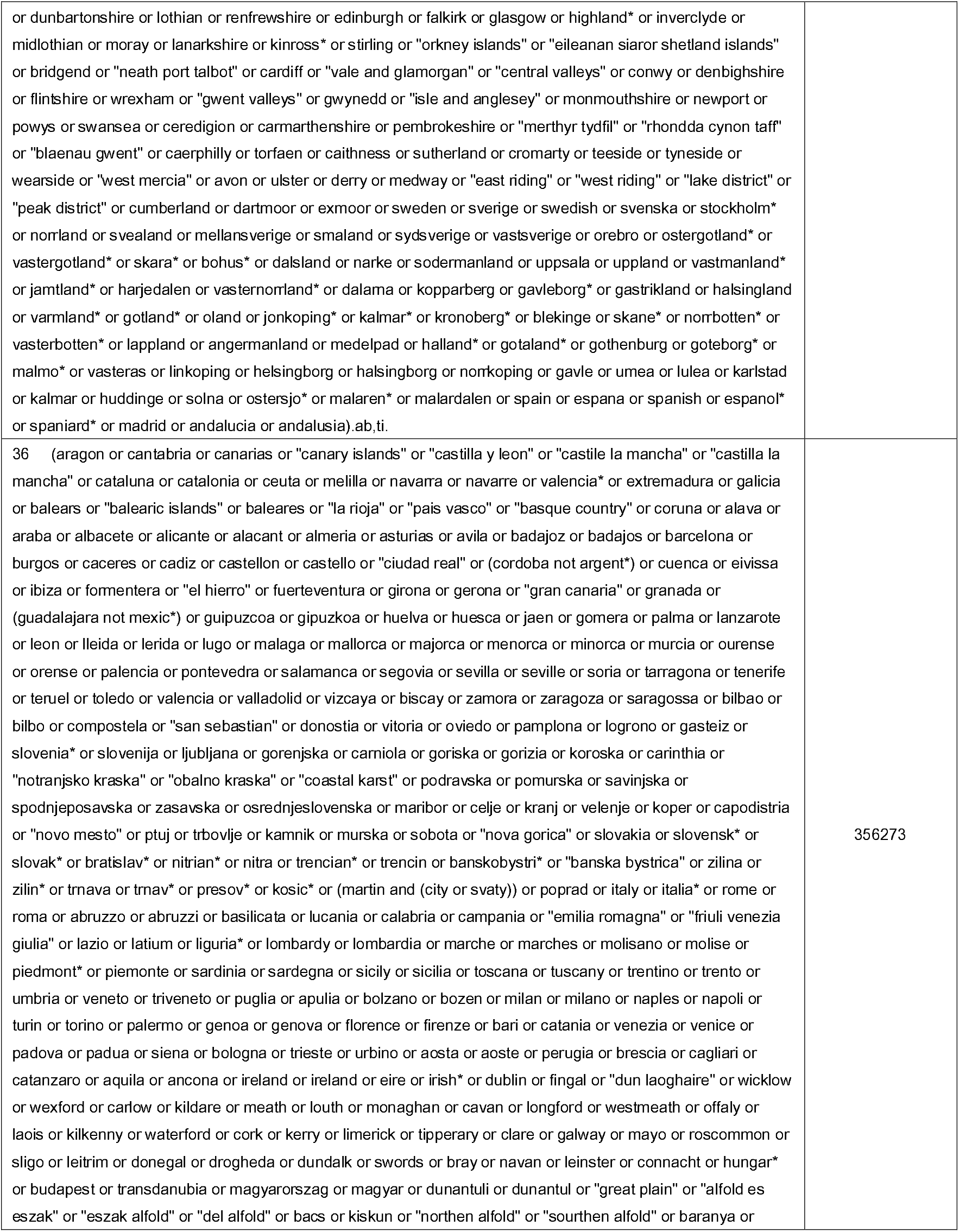
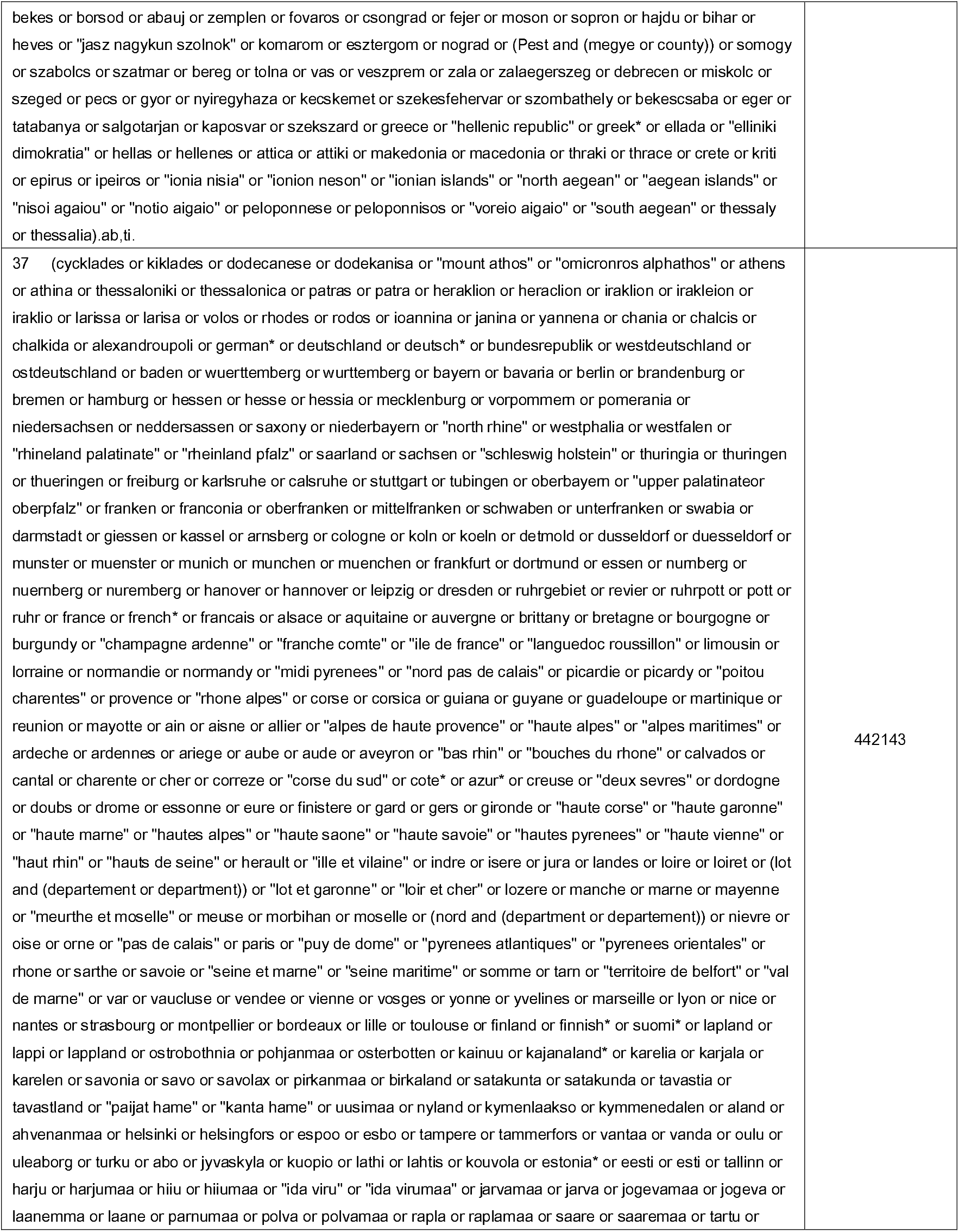
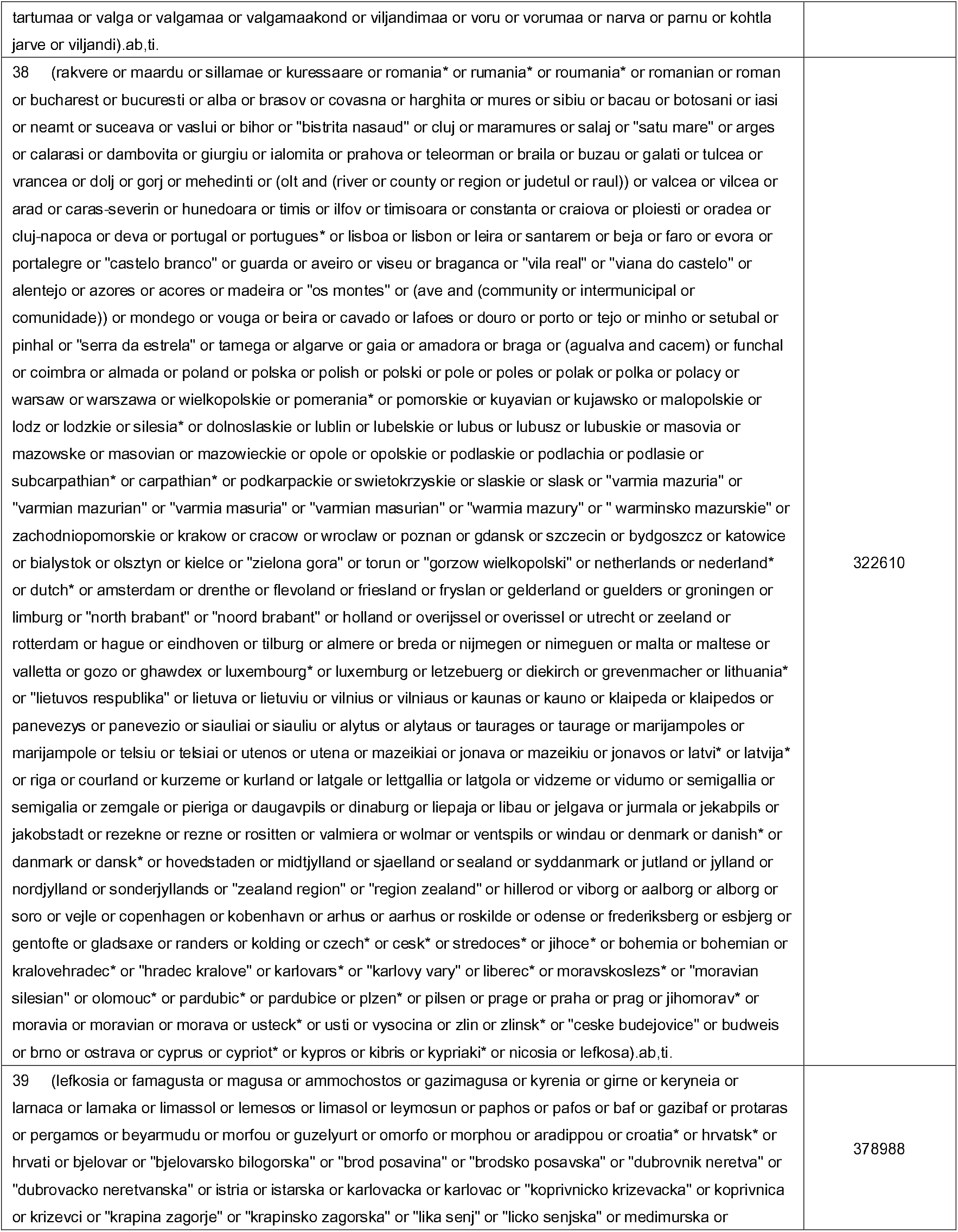
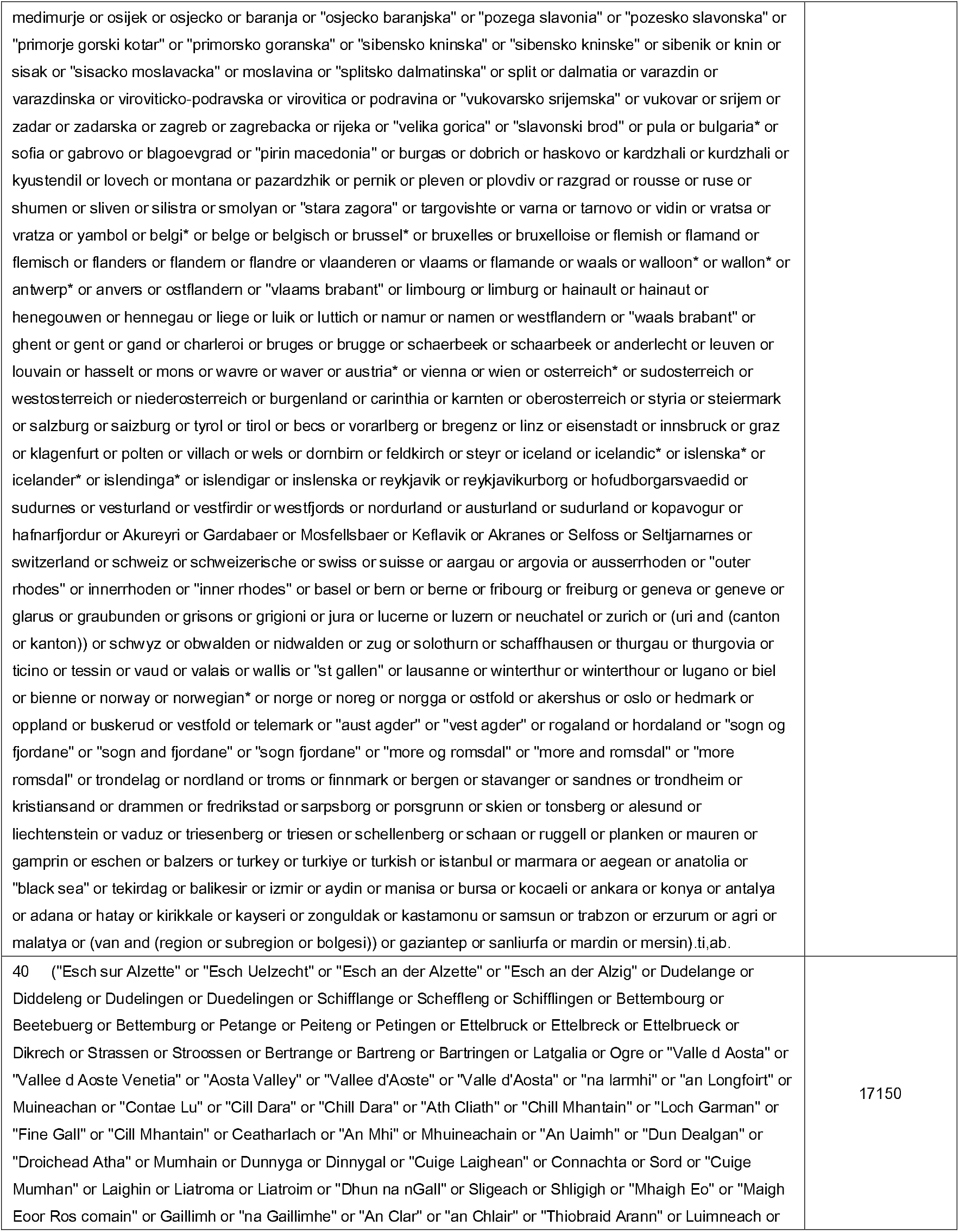
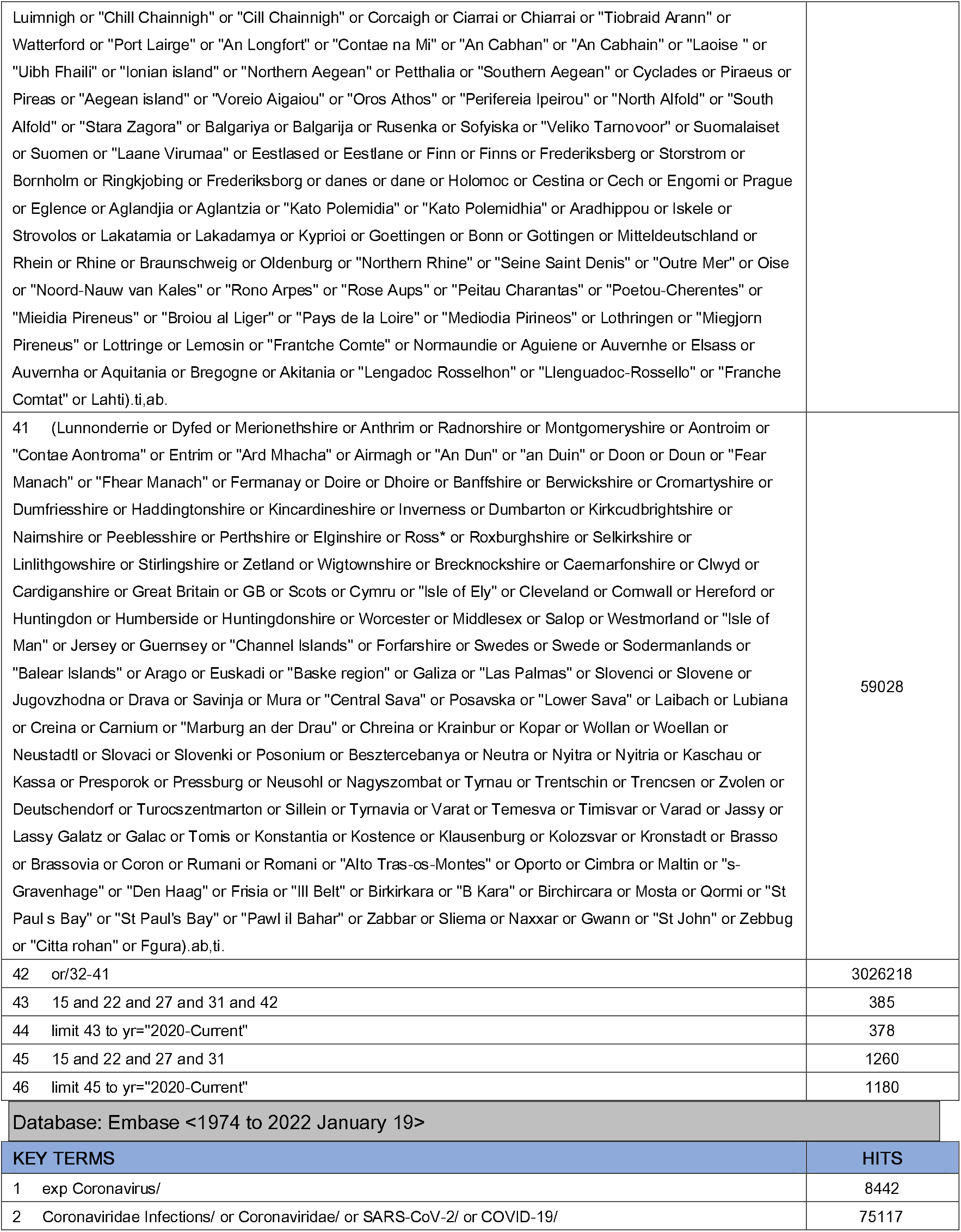
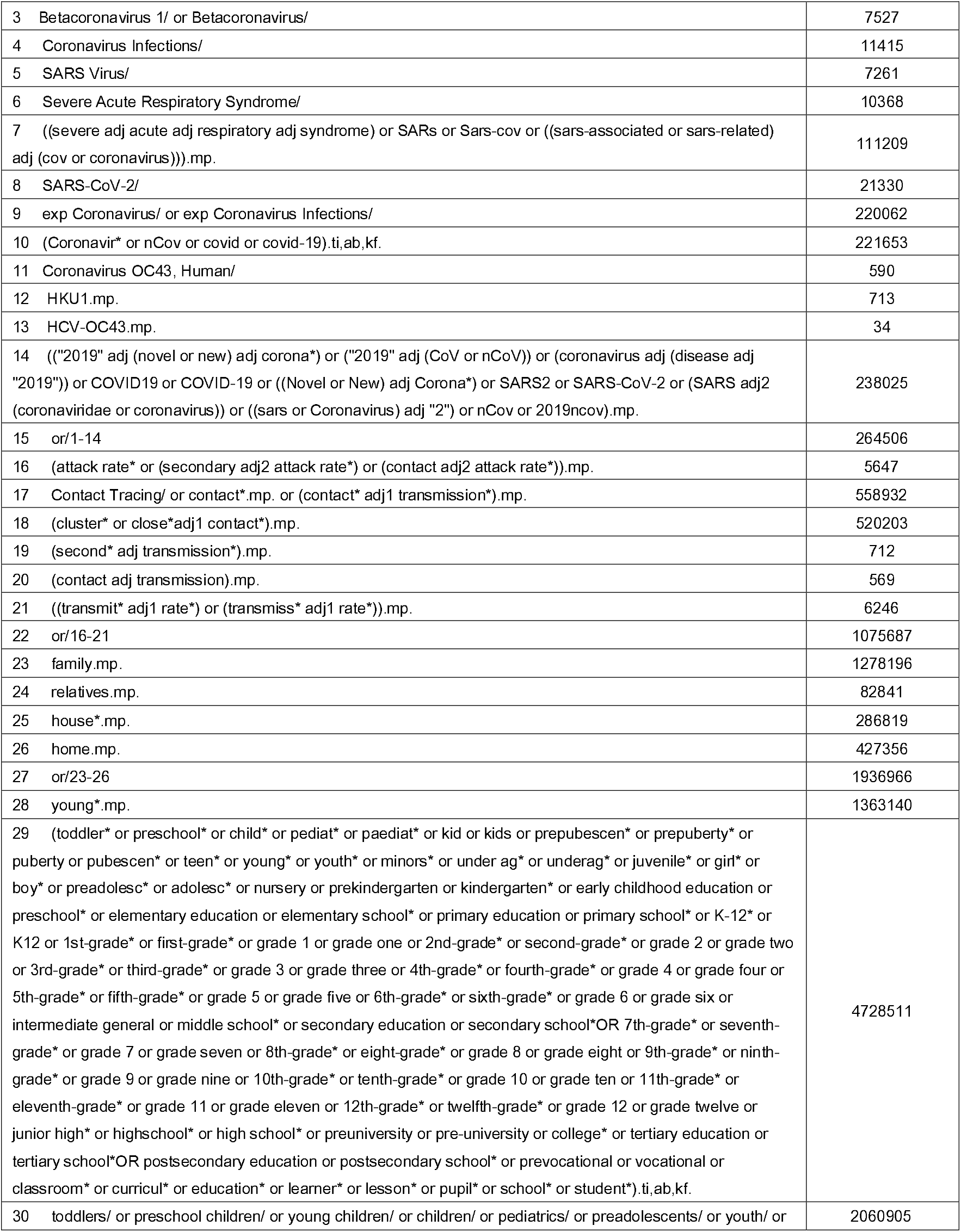
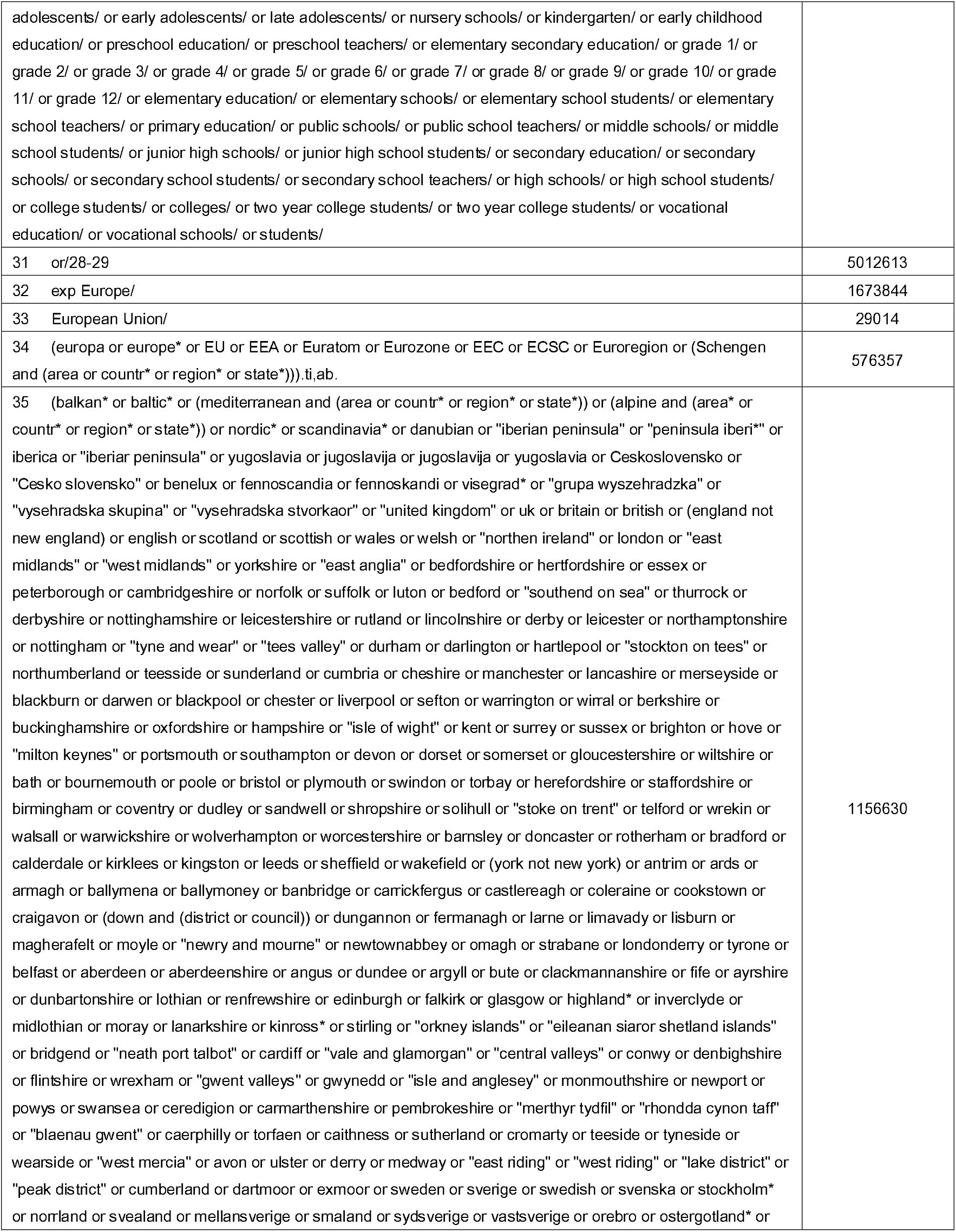
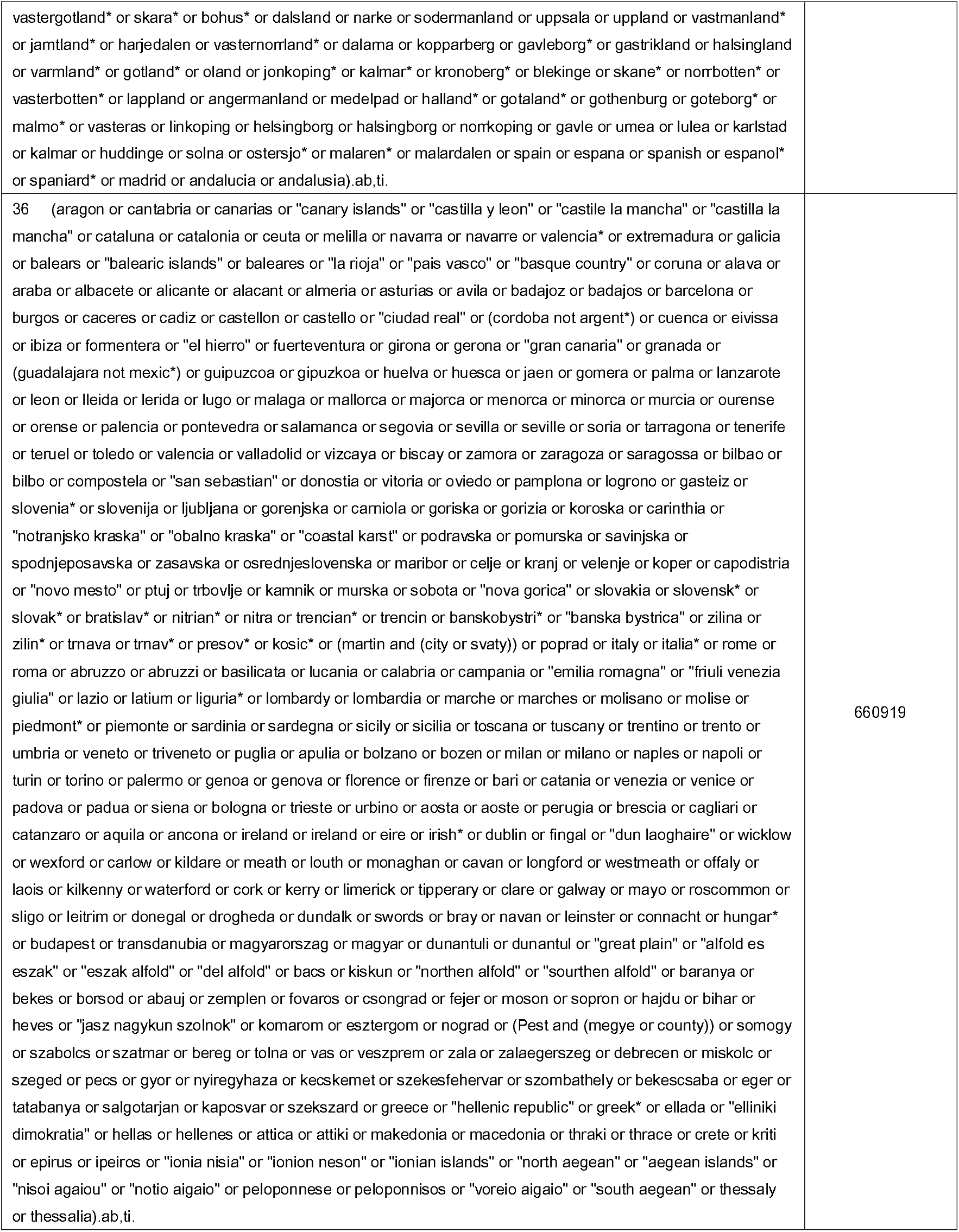
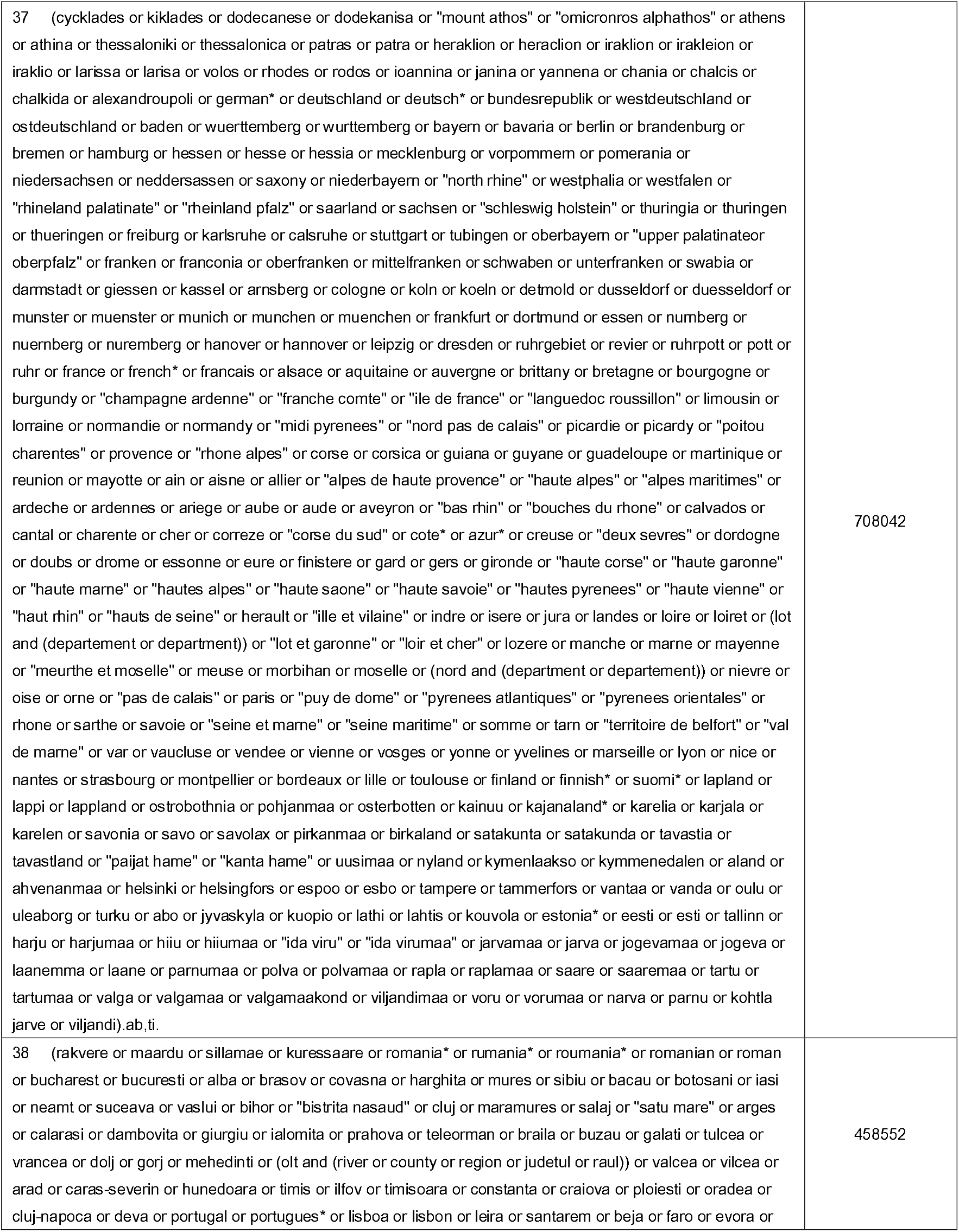
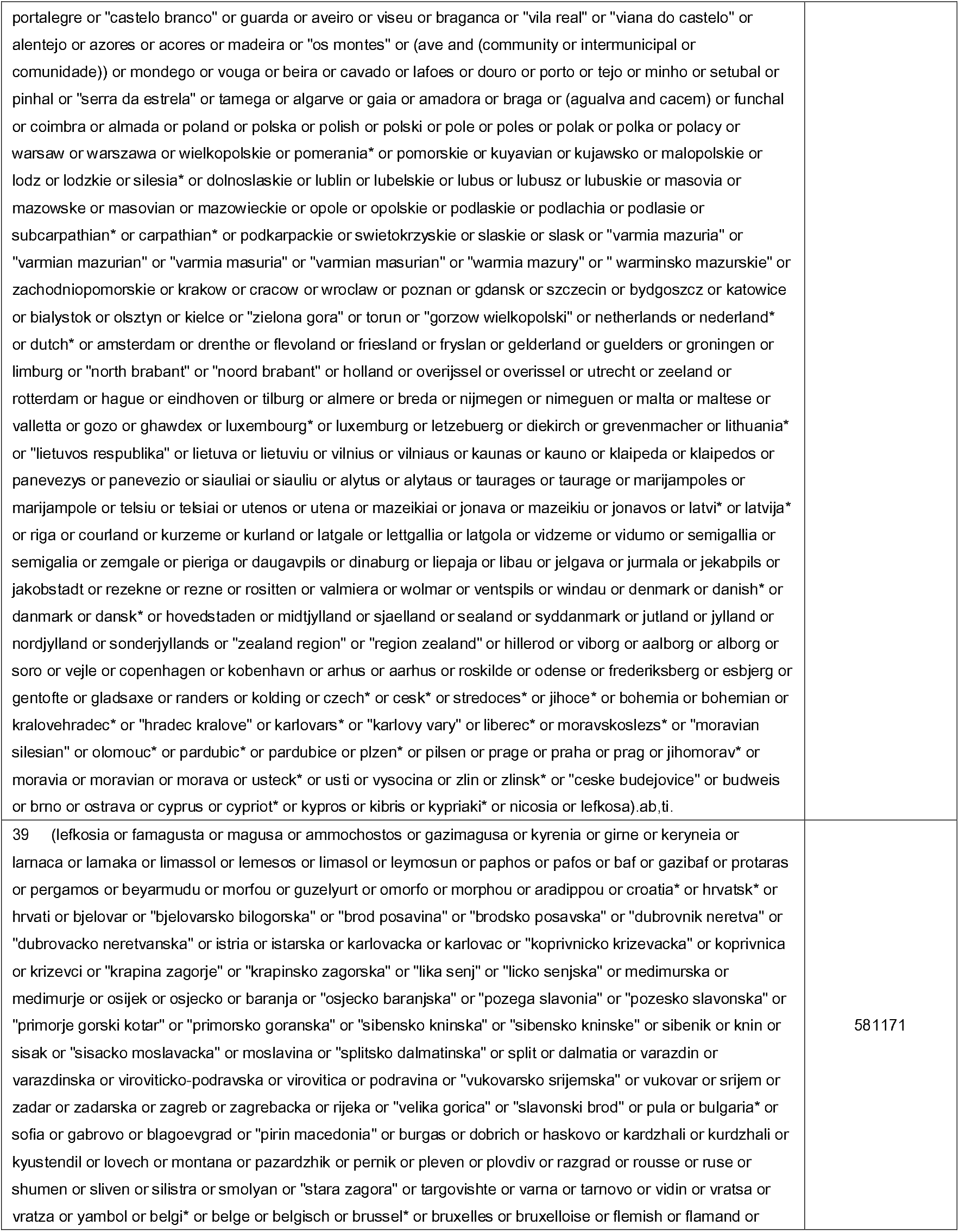
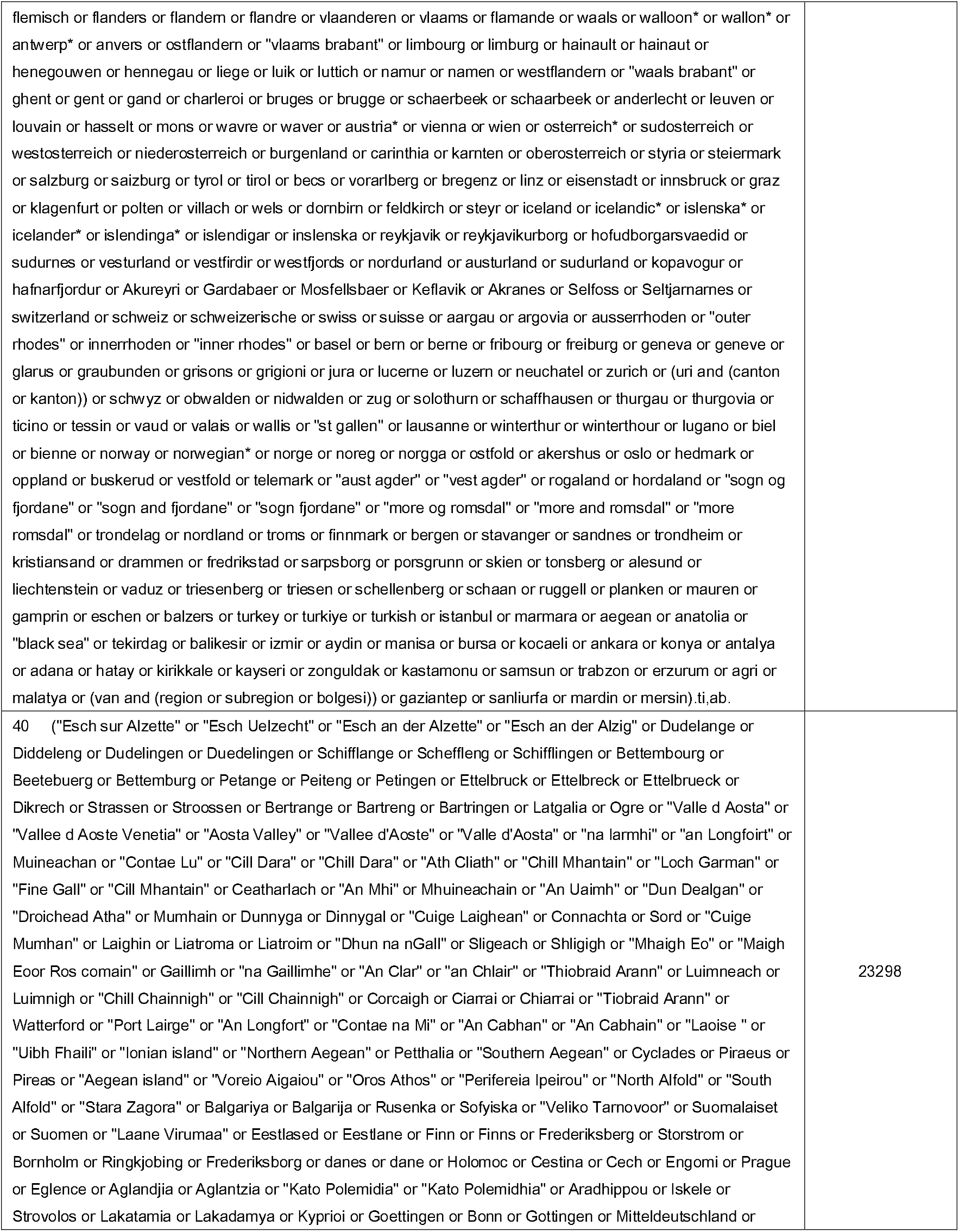
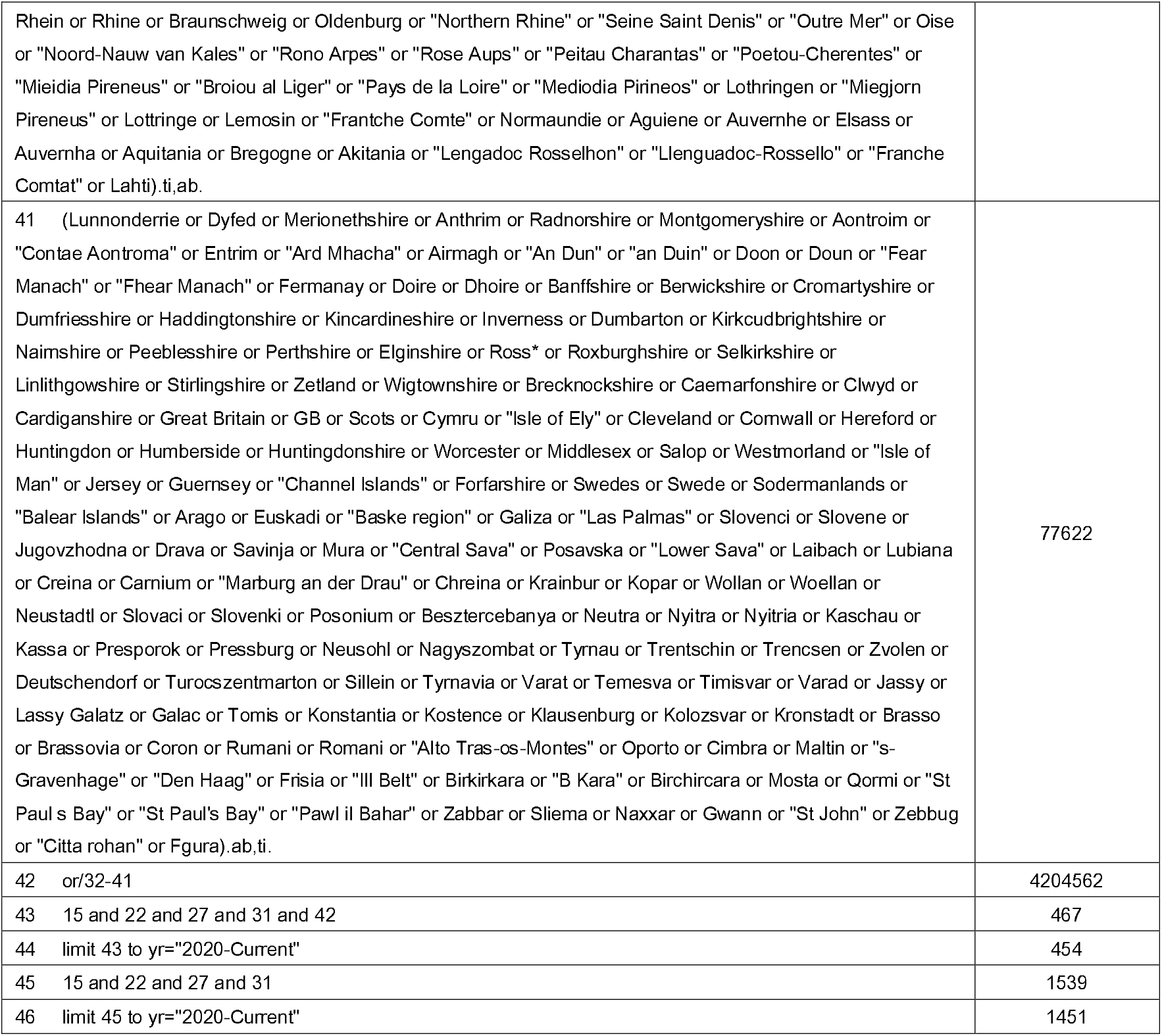
Search strategies for identifying studies

**Supplementary Material****Supplementary Table 1: Search strategies for identifying studies** Database: Ovid MEDLINE(R) ALL <1946 to January 19, 2022>
| KEY TERMS | HITS |
| --- | --- |
| 1 exp Coronavirus/ | 4887 |
| 2 Coronaviridae Infections/ or Coronaviridae/ or SARS-CoV-2/ or COVID-19/ | 137004 |
| 3 Betacoronavirus 1/ or Betacoronavirus/ | 33283 |
| 4 Coronavirus Infections/ | 45290 |
| 5 SARS Virus/ | 4007 |
| 6 Severe Acute Respiratory Syndrome/ | 5658 |
| 7 ((severe adj acute adj respiratory adj syndrome) or SARs or Sars-cov or ((sars-associated or sars-related) adj (cov or coronavirus))).mp. | 152979 |
| 8 SARS-CoV-2/ | 105907 |
| 9 exp Coronavirus/ or exp Coronavirus Infections/ | 151122 |
| 10 (Coronavir* or nCov or covid or covid-19).ti,ab,kf. | 216182 |
| 11 Coronavirus OC43, Human/ | 269 |
| 12 HKU1.mp. | 490 |
| 13 HCV-OC43.mp. | 37 |
| 14 (("2019" adj (novel or new) adj corona*) or ("2019" adj (CoV or nCoV)) or (coronavirus adj (disease adj "2019")) or COVID19 or COVID-19 or ((Novel or New) adj Corona*) or SARS2 or SARS-CoV-2 or (SARS adj2 (coronaviridae or coronavirus)) or ((sars or Coronavirus) adj "2") or nCov or 2019ncov).mp. | 221107 |
| 15 or/1-14 | 244428 |
| 16 (attack rate* or (secondary adj2 attack rate*) or (contact adj2 attack rate*)).mp. | 4942 |
| 17 Contact Tracing/ or contact*.mp. or (contact* adj1 transmission*).mp. | 451220 |
| 18 (cluster* or close*adj1 contact*).mp. | 452890 |
| 19 (second* adj transmission*).mp. | 630 |
| 20 (contact adj transmission).mp. | 500 |
| 21 ((transmit* adj1 rate*) or (transmiss* adj1 rate*)).mp. | 5830 |
| 22 or/16-21 | 901306 |
| 23 family.mp. | 1084846 |
| 24 relatives.mp. | 63350 |
| 25 house*.mp. | 215827 |
| 26 home.mp. | 277244 |
| 27 or/23-26 | 1527765 |
| 28 young*.mp. | 1611781 |
| 29 (toddler* or preschool* or child* or pediat* or paediat* or kid or kids or prepubescen* or prepuberty* or puberty or pubescen* or teen* or young* or youth* or minors* or under ag* or underag* or juvenile* or girl* or boy* or preadolesc* or adolesc* or nursery or prekindergarten or kindergarten* or early childhood education or preschool* or elementary education or elementary school* or primary education or primary school* or K-12* or K12 or 1st-grade* or first-grade* or grade 1 or grade one or 2nd-grade* or second-grade* or grade 2 or grade two or 3rd-grade* or third-grade* or grade 3 or grade three or 4th-grade* or fourth-grade* or grade 4 or grade four or | 3665192 |
| 5th-grade* or fifth-grade* or grade 5 or grade five or 6th-grade* or sixth-grade* or grade 6 or grade six or intermediate general or middle school* or secondary education or secondary school*OR 7th-grade* or seventh-grade* or grade 7 or grade seven or 8th-grade* or eight-grade* or grade 8 or grade eight or 9th-grade* or ninth-grade* or grade 9 or grade nine or 10th-grade* or tenth-grade* or grade 10 or grade ten or 11th-grade* or eleventh-grade* or grade 11 or grade eleven or 12th-grade* or twelfth-grade* or grade 12 or grade twelve or junior high* or highschool* or high school* or preuniversity or pre-university or college* or tertiary education or tertiary school*OR postsecondary education or postsecondary school* or prevocational or vocational or classroom* or curricular* or education* or learner* or lesson* or pupil* or school* or student*).ti,ab,kf. |  |
| 30 toddlers/ or preschool children/ or young children/ or children/ or pediatrics/ or preadolescents/ or youth/ or adolescents/ or early adolescents/ or late adolescents/ or nursery schools/ or kindergarten/ or early childhood education/ or preschool education/ or preschool teachers/ or elementary secondary education/ or grade 1/ or grade 2/ or grade 3/ or grade 4/ or grade 5/ or grade 6/ or grade 7/ or grade 8/ or grade 9/ or grade 10/ or grade 11/ or grade 12/ or elementary education/ or elementary schools/ or elementary school students/ or elementary school teachers/ or primary education/ or public schools/ or public school teachers/ or middle schools/ or middle school students/ or junior high schools/ or junior high school students/ or secondary education/ or secondary schools/ or secondary school students/ or secondary school teachers/ or high schools/ or high school students/ or college students/ or colleges/ or two year college students/ or two year college students/ or vocational education/ or vocational schools/ or students/ | 3296611 |
| 31 or/28-29 | 4355387 |
| 32 exp Europe/ | 1507261 |
| 33 European Union/ | 17061 |
| 34 (europa or europe* or EU or EEA or Euratom or Eurozone or EEC or ECSC or Euroregion or (Schengen and (area or countr* or region* or state*))).ti,ab. | 347879 |
| 35 (balkan* or baltic* or (mediterranean and (area or countr* or region* or state*)) or (alpine and (area* or countr* or region* or state*)) or nordic* or scandinavia* or danubian or "iberian peninsula" or "peninsula iberi*" or iberica or "iberiar peninsula" or yugoslavia or jugoslavija or jugoslavija or yugoslavia or Ceskoslovensko or "Cesko slovensko" or benelux or fennoscandia or fennoskandi or visegrad* or "grupa wyszehradzka" or "vysehradzka skupina" or "vysehradzka stvorkaor" or "united kingdom" or uk or britain or british or (england not new england) or english or scotland or scottish or wales or welsh or "northern ireland" or london or "east midlands" or "west midlands" or yorkshire or "east anglia" or bedfordshire or hertfordshire or essex or peterborough or cambridgeshire or norfolk or suffolk or luton or bedford or "southend on sea" or thurrock or derbyshire or nottinghamshire or leicestershire or rutland or lincolnshire or derby or leicester or northamptonshire or nottingham or "tyne and wear" or "tees valley" or durham or darlington or hartlepool or "stockton on tees" or northumberland or teesside or sunderland or cumbria or cheshire or manchester or lancashire or merseyside or blackburn or darwen or blackpool or chester or liverpool or sefton or warrington or wirral or berkshire or buckinghamshire or oxfordshire or hampshire or "isle of wight" or kent or surrey or sussex or brighton or hove or "milton keynes" or portsmouth or southampton or devon or dorset or somerset or gloucestershire or wiltshire or bath or bournemouth or poole or bristol or plymouth or swindon or torbay or herefordshire or staffordshire or birmingham or coventry or dudley or sandwell or shropshire or solihull or "stoke on trent" or telford or wrekin or walsall or warwickshire or wolverhampton or worcestershire or barnsley or doncaster or rotherham or bradford or calderdale or kirklees or kingston or leeds or sheffield or wakefield or (york not new york) or antrim or ards or armagh or ballymena or ballymoney or banbridge or carrickfergus or castlereagh or coleraine or cookstown or craigavon or (down and (district or council)) or dungannon or fermanagh or larne or limavady or lisburn or magherafelt or moyle or "newry and mourne" or newtownabbey or omagh or strabane or londonderry or tyrone or belfast or aberdeen or aberdeenshire or angus or dundee or argyll or bute or clackmannanshire or fife or ayrshire | 783596 |
| or dunbartonshire or lothian or renfrewshire or edinburgh or falkirk or glasgow or highland* or inverclyde or midlothian or moray or lanarkshire or kinross* or stirling or "orkney islands" or "eileanan siaror shetland islands" or bridgend or "neath port talbot" or cardiff or "vale and glamorgan" or "central valleys" or conwy or denbighshire or flintshire or wrexham or "gwent valleys" or gwynedd or "isle and anglesey" or monmouthshire or newport or powys or swansea or ceredigion or carmarthenshire or pembrokeshire or "merthyr tydfil" or "rhondda cynon taff" or "blaenau gwent" or caerphilly or torfaen or caithness or sutherland or cromarty or teeside or tyneside or wearside or "west mercia" or avon or ulster or derry or midway or "east riding" or "west riding" or "lake district" or "peak district" or cumberland or dartmoor or exmoor or sweden or sverige or swedish or svenska or stockholm* or norrland or svealand or mellansverige or smaland or sydsverige or vastsverige or orebro or ostergotland* or vastergotland* or skara* or bohus* or dalsland or narke or sodermanland or uppsala or uppland or vastmanland* or jamtland* or harjedalen or vasternorrland* or dalarna or kopparberg or gavleborg* or gastrikland or halsingland or varmland* or gotland* or oland or jonkoping* or kalmars* or kronoberg* or blekinge or skane* or norrbotten* or vasterbotten* or lappland or angermanland or medelpad or halland* or gotaland* or gothenburg or goteborg* or malmo* or vasteras or linkoping or helsingborg or halsingborg or norrkoping or gavle or umea or lulea or karlstad or kalmar or huddinge or solna or ostersjo* or malaren* or malardalen or spain or espana or spanish or espanol* or spaniard* or madrid or andalucia or andalusia) ab,ti. |  |
| 36 (aragon or cantabria or canarias or "canary islands" or "castilla y leon" or "castile la mancha" or "castilla la mancha" or cataluna or catalonia or ceuta or melilla or navarra or navarre or valencia* or extremadura or galicia or balears or "balearic islands" or baleares or "la rioja" or "pais vasco" or "basque country" or coruna or alava or araba or albacete or alicante or alacant or almeria or asturias or avila or badajoz or badajos or barcelona or burgos or caceres or cadiz or castellon or castello or "ciudad real" or (cordoba not argent*) or cuenca or eivissa or ibiza or formentera or "el hierro" or fuerteventura or girona or gerona or "gran canaria" or granada or (guadalajara not mexic*) or guipuzcoa or gipuzkoa or huelva or huesca or jaen or gomera or palma or lanzarote or leon or lleida or lerida or lugo or malaga or mallorca or majorca or menorca or minorca or murcia or ourense or orense or palencia or pontevedra or salamanca or segovia or sevilla or seville or soria or tarragona or tenerife or teruel or toledo or valencia or valladolid or vizcaya or biscay or zamora or zaragoza or saragossa or bilbao or bilbo or compostela or "san sebastian" or donostia or vitoria or oviedo or pamplona or logrono or gasteiz or slovenia* or slovenija or ljubljana or gorenjska or carniola or goriska or gorizia or koroska or carinthia or "notranjsko kraska" or "obalno kraska" or "coastal karst" or podravska or pomurska or savinjska or spodnjeposavska or zasavska or osrednjeslovenska or maribor or celje or kranj or velenje or koper or capodistria or "novo mesto" or ptuj or trbovlje or kamnik or murska or sobota or "nova gorica" or slovakia or slovensk* or slovak* or bratislav* or nitria* or nitra or trencian* or trencin or banksobystr* or "banska bystrica" or zilina or zilin* or trnava or trnav* or presov* or kosic* or (martin and (city or svaty)) or poprad or italy or italia* or rome or roma or abruzzo or abruzz* or basilicata or lucania or calabria or campania or "emilia romagna" or "friuli venezia giulia" or lazio or latium or liguria* or lombardy or lombardia or marche or marches or molisano or molise or piedmont* or piemonte or sardinia or sardegna or sicily or sicilia or toscana or tuscany or trentino or trento or umbria or veneto or triveneto or puglia or apulia or bolzano or bozen or milan or milano or naples or napoli or turin or torino or palermo or genoa or genova or florence or firenze or bari or catania or venezia or venice or padova or padua or siena or bologna or trieste or urbino or aosta or aoste or perugia or brescia or cagliari or catanzaro or aquila or ancona or ireland or ireland or eire or irish* or dublin or fingsal or "dun laoghaire" or wicklow or wexford or carlow or kildare or meath or louth or monaghan or cavan or longford or westmeath or offaly or laois or kilkenny or waterford or cork or kerry or limerick or tipperary or clare or galway or mayo or roscommon or sligo or leitrim or donegal or drogheda or dundalk or swords or bray or navan or leinster or connacht or hungar* or budapest or transdanubia or magyarország or magyar or dunantuli or dunantul or "great plain" or "alföld es eszak" or "eszak alföld" or "del alföld" or bacs or kiskun or "northern alföld" or "southern alföld" or baranya or | 356273 |
| bekes or borsod or abauj or zemplen or fovaros or csongrad or fejer or moson or sopron or hajdu or bihar or heves or "jasz nagykun szolnok" or komarom or esztergom or nograd or (Pest and (megye or county)) or somogy or szabolcs or szatmar or bereg or toina or vas or veszprem or zala or zalaegerszeg or debrecen or miskolc or szeged or pecs or gyor or nyiregyhaza or kecskemet or szekesfehervar or szombathely or bekescsaba or eger or tatabanya or salgotarjan or kaposvar or szekszard or greece or "hellenic republic" or greek* or ellada or "elliniki dimokratia" or hellas or hellenes or attica or attiki or makedonia or macedonia or thraki or thrace or crete or kriti or epirus or ipeiros or "ionia nisia" or "ionion neson" or "ionian islands" or "north aegean" or "aegean islands" or "nisoï agaiou" or "notio aigaiο" or peloponnese or peloponnisos or "voreio aigaiο" or "south aegean" or thessaly or thessalia).ab.ti. |  |
| 37 (cycklades or kijklades or dodecanese or dodekanisa or "mount athos" or "omicronros alphathos" or athens or athina or thessaloniki or thessalonica or patras or patra or heraklion or heraclion or iraklion or irakleion or iraklio or larissa or larisa or volos or rhodes or rodos or ioannina or janina or yannena or chania or chalcis or chalkida or alexandroupoli or german* or deutschland or deutsch* or bundesrepublik or westdeutschland or ostdeutschland or baden or wuerttemberg or wurtemberg or bayern or bavaria or berlin or brandenburg or bremen or hamburg or hessen or hesse or hessia or mecklenburg or vorpommern or pomerania or niedersachsen or neddersassen or saxony or niederbayern or "north rhine" or westphalia or westfalen or "rhineland palatinate" or "rheinland pfalz" or saarland or sachsen or "schleswig holstein" or thuringia or thuringen or thueringen or freiburg or karlsruhe or callsruhe or stuttgart or tuingen or oberbayern or "upper palatinate" or oberpfalz" or franken or franconia or oberfranken or mittelfranken or schwaben or unterfranken or swabia or darmstadt or giessen or kassel or arnsberg or cologne or koln or koeln or detmold or dusseldorf or duesseldorf or munster or muenster or munich or munchen or muenchen or frankfurt or dortmund or essen or numberg or nuernberg or nuremberg or hanover or hannover or leipzig or dresden or ruhrgebiet or revier or ruhrpott or pott or ruhr or france or french* or francais or alsace or aquitaine or auvergne or brittany or bretagne or bourgogne or burgundy or "champagne ardenne" or "franche comte" or "ile de france" or "languedoc roussillon" or limousin or lorraine or normandie or normandy or "midi pyrenees" or "nord pas de calais" or picardie or picardy or "poitou charentes" or provence or "rhone alpes" or corse or corsica or guiana or guyane or guadeloupe or martinique or reunion or mayotte or ain or aise or allier or "alpes de haute provence" or "haute alpes" or "alpes maritimes" or ardeche or ardennes or ariege or aube or aude or aveyron or "bas rhin" or "bouches du rhone" or calvados or cantal or charente or cher or correze or "corse du sud" or cote* or azur* or creuse or "deux sevrès" or dordogne or doubs or drome or essonne or eure or finistere or gard or gers or gironde or "haute corse" or "haute garonne" or "haute marne" or "hautes alpes" or "haute saone" or "haute savoie" or "hautes pyrenees" or "haute vienne" or "haut rhin" or "hauts de seine" or herault or "ille et vilaine" or indre or isere or jura or landes or loire or loiret or (lot and (departement or department)) or "lot et garonne" or "loir et cher" or lozere or manche or marne or mayenne or "meurthe et moselle" or meuse or morbihan or moselle or (nord and (department or departement)) or nievre or oise or orne or "pas de calais" or paris or "puy de dome" or "pyrenees atlantiques" or "pyrenees orientales" or rhone or sarthe or savoie or "seine et marne" or "seine maritime" or somme or tarn or "territoire de belfort" or "val de marne" or var or vaucluse or vendee or vienne or vosges or yonne or yvelines or marseille or lyon or nice or nantes or strasbourg or montpellier or bordeaux or lille or toulouse or finland or finnish* or suomi* or lapland or lappi or lappland or ostrobothnia or pohjanmaa or osterbotten or kainuu or kajanaland* or karelia or karjala or karelen or savonia or savo or savolax or pirkanmaa or birkaland or satakunta or satakunda or tavastia or tavastland or "pajjat hame" or "kanta hame" or uusimaa or nyland or kymenlaakso or kymmenedalen or aland or ahvenanmaa or helsinki or helsingfors or espoo or esbo or tampere or tammerfors or vantaa or vanda or oulu or uleaborg or turku or abo or jyvaskyla or kuopio or lathi or lahtis or kouvola or estonia* or eesti or esti or tallinn or harju or harjumaa or hiiu or hiiumaa or "ida viru" or "ida virumaa" or jarvamaa or jarva or jogevamaa or jogeva or laanemaa or laane or parnumaa or polva or polvamaa or rapla or raplamaa or saare or saaremaa or tartu or | 442143 |
| tartumaa or valga or valgamaa or valgamaakond or viljandimaa or voru or vorumaa or narva or parnu or kohtla jarve or viljandi).ab,ti. |  |
| 38 (rakvere or maardu or sillamae or kuressaare or romania* or rumania* or roumania* or romanian or roman or bucharest or Bucuresti or alba or brasov or covasna or harghita or mures or sibiu or bacau or botosani or iasi or neamt or suceava or vaslui or bihor or "bistrita nasaud" or cluj or maramures or salaj or "satu mare" or arges or calarasi or dambovitza or giurgiu or ialomita or prahova or teleorman or braila or buzau or galati or tulcea or vrancea or dolj or gorj or mehedinti or (olt and (river or county or region or judetul or raul)) or valcea or vilcea or arad or caras-severin or hunedoara or timis or ilfov or timisoara or constanta or craiova or ploiesti or oradea or cluj-napoca or deva or portugal or portugues* or lisboa or lisbon or leira or santarem or beja or faro or evora or portalegre or "castelo branco" or guarda or aveiro or viseu or braganca or "vila real" or "viana do castelo" or alentejo or azores or acores or madeira or "os montes" or (ave and (community or intermunicipal or comunidade)) or mondego or vouga or beira or cavado or lafoes or douro or porto or tejo or minho or setubal or pinhal or "serra da estrela" or tamega or algarve or gaia or amadora or braga or (agualva and cacem) or funchal or coimbra or almada or poland or polska or polish or polski or pole or poles or polak or polka or polacy or warsaw or warszawa or wielkopolskie or pomerania* or pomorskie or kujavian or kujawsko or malopolskie or lodz or lodzkie or silesia* or dolnoslaskie or lublin or lubelskie or lubus or lubusz or lubuskie or masovia or mazowske or masovian or mazowieckie or opole or opolskie or podlaskie or podlachia or podlasie or subcarpathian* or carpathian* or podkarpackie or swietokrzyskie or slaskie or slask or "varmia mazuria" or "varmian mazurian" or "varmia masuria" or "varmian masurian" or "warmia mazury" or "warminsko mazurskie" or zachodniopomorskie or krakow or cracow or wroclaw or poznan or gdansk or szczecin or bydgoszcz or katowice or bialystok or olsztyn or kielce or "zielona gora" or torun or "gorzow wielkopolski" or netherlands or nederland* or dutch* or amsterdam or drenthe or flevoland or friesland or frysian or gelderland or guelders or groningen or limburg or "north brabant" or "noord brabant" or holland or overijssel or overijssel or utrecht or zeeland or rotterdam or hague or eindhoven or tilburg or almere or breda or nijmegen or nijmegen or malta or maltese or valletta or gozo or ghawdex or luxembourg* or luxemburg or letzebuerg or diekirch or grevenmacher or lithuania* or "lietuvos respublika" or lietuva or lietuvii or vilnius or vilniaus or kaunas or kauno or klaipeda or klaipedos or panevezys or panevezio or siauliai or siauliu or alytus or alytaus or taurages or taurage or marijampoles or marijampole or telsiu or telsiai or utenas or utena or mazeikiai or jonava or mazeikiai or jonavos or latvi* or latvija* or riga or courland or kurzeme or kurland or latgale or lettigallia or latgola or vidzeme or vidumo or semigallia or semigalia or zemgale or pieriga or daugavpils or dinaburg or iepaja or libau or jelgava or jurmala or jekabpils or jakobstadt or rezekne or rezne or rositten or valmiera or wolmar or ventspils or windau or denmark or danish* or danmark or dansk* or hovedstaden or midtjylland or sjælland or sealand or syddanmark or jutland or jylland or nordjylland or sønderjyllands or "zealand region" or "region zealand" or hillerød or viborg or aalborg or alborg or soro or vejle or copenhagen or københavn or århus or aarhus or roskilde or odense or frederiksberg or esbjerg or gentofte or gladsaxe or randers or kolding or czech* or cesk* or stredoces* or jihoce* or bohemia or bohemian or kralovehradec* or "hradec kralove" or karlovars* or "karlovy vary" or liberec* or moravskoslezs* or "moravian silesian" or olomouc* or pardubice* or pardubice or plzen* or pilsen or prague or praha or prag or jihomorav* or moravia or moravian or morava or usteck* or usti or vysocina or zlin or zlíns* or "ceske budejovice" or budweis or brno or ostrava or cyprus or cypriot* or kypros or kibris or kypraki* or nicosia or lefkosa).ab,ti. | 322610 |
| 39 (lefkosia or famagusta or magusa or ammochoostos or gazimagusa or kyrenia or girne or keryneia or larnaca or larnaka or limassol or lemesos or limasol or leymosun or paphos or pafos or baf or gazibaf or protaras or pergamos or beyarmudu or morfou or guzelyurt or omorfo or morphou or aradippou or croatia* or hrvatsk* or hrvati or bjelovar or "bjelovarsko bilogorska" or "brod posavina" or "brodsko posavska" or "dubrovnik neretva" or "dubrovacko neretvanska" or istria or istarska or karlovacka or karlovac or "kopriivnicko krizevacka" or kopriivnica or krizevci or "krapina zagorje" or "krapinsko zagorska" or "lika senj" or "licko senjska" or medimurska or | 378988 |
| medimurje or osijek or osjecko or baranja or "osjecko baranjska" or "pozega slavonia" or "pozesko slavonska" or "primorje gorski kotar" or "primorsko goranska" or "sibensko kninska" or "sibensko kninske" or sibenik or knin or sisak or "sisacko moslavacka" or moslavina or "splitsko dalmatinska" or split or dalmatia or varazdin or varazdinska or viroviticko-podravskaa or virovitica or podravina or "vukovarsko srijemska" or vukovar or srijem or zadar or zadarska or zagreb or zagrebacka or rijeka or "velika gorica" or "slavonski brod" or pula or bulgaria* or sofia or gabrovo or blagoevgrad or "pirin macedonia" or burgas or dobrich or haskovo or kardzhali or kurdzhali or kyustendil or lovech or montana or pazardzhik or pernik or pleven or plovdiv or razgrad or rousse or ruse or shumen or sliven or silistra or smolyan or "stara zagora" or targovishte or varna or tarnovo or vidin or vratsa or vratza or yambol or belgi* or belge or belgisch or brussel* or bruxelles or bruxelloise or flemish or flamand or flemisch or flanders or flandern or flandre or vlaanderen or vlaams or flamande or waals or walloon* or wallon* or antwerp* or anvers or ostflandern or "vlaams brabant" or limburg or limburg or hainault or hainaut or henegouwen or hennegau or liege or luik or luttich or namur or namen or westflandern or "waals brabant" or ghent or gent or gand or charleroi or bruges or brugge or schaerbeek or schaarbeek or anderlecht or leuven or louvain or hasselt or mons or wavre or waver or austria* or vienna or wien or osterreich* or sudosterreich or westosterreich or niederosterreich or burgenland or carinthia or karnten or oberosterreich or styria or steiermark or salzburg or saizburg or tyrol or tirol or becs or vorarlberg or bregenz or linz or eisenstadt or innsbruck or graz or klagenfurt or polten or villach or wels or dornbirn or feldkirch or steyr or iceland or icelandic* or islenska* or icelander* or islendinga* or islendigar or inslenska or reykjavik or reykjavikurborg or hofudborgarsvaedid or sudurnes or vesturland or vestfirðir or westfjords or nordurland or austurland or sudurland or kopavogur or hafnarfjörður or Akureyri or Gardabaer or Mosfellsbaer or Keflavik or Akranes or Selfoss or Seltjarnarnes or switzerland or schweiz or schweizerische or swiss or suisse or aargau or argovia or ausser rhoden or "outer rhodes" or innerrhoden or "inner rhodes" or basel or bern or berne or fribourg or freiburg or geneva or geneve or glarus or graubunden or grisons or grigioni or jura or lucerne or luzern or neuchatel or zurich or (uri and (canton or kanton)) or schwyz or obwalden or nidwalden or zug or solothurn or schaffhausen or thurgau or thurgovia or ticino or tessin or vaud or valais or wallis or "st gallen" or lausanne or winterthur or winterthour or lugano or biel or bienne or norway or norwegian* or norge or noreg or norgga or ostfold or akershus or oslo or hedmark or oppland or buskerud or vestfold or telemark or "aust agder" or "vest agder" or rogaland or hordaland or "sogn og fjordane" or "sogn and fjordane" or "sogn fjordane" or "more og romsdal" or "more and romsdal" or "more romsdal" or trondelag or nordland or troms or finnmark or bergen or stavanger or sandnes or trondheim or kristiansand or drammen or fredrikstad or sarpsborg or porsgrunn or skien or tonsberg or alesund or liechtenstein or vaduz or triesenberg or triesen or schellenberg or schaan or ruggell or planken or mauren or gamprin or eschen or balzers or turkey or turkiye or turkish or istanbul or marmara or aegean or anatolia or "black sea" or tekirdag or balikesir or izmir or aydin or manisa or bursa or kocaeli or ankara or konya or antalya or adana or hatay or kirikkale or kayseri or zonguldak or kastamonu or samsun or trabzon or erzurum or agri or malatya or (van and (region or subregion or bolgesi)) or gaziantep or sanliurfa or mardin or mersin).ti.ab. |  |
| 40 ("Esch sur Alzette" or "Esch Uelzecht" or "Esch an der Alzette" or "Esch an der Alzig" or Dudelange or Diddeleng or Dudelingen or Duedelingen or Schiffange or Scheffleng or Schifflingen or Bettembourg or Beetebeuerg or Bettemburg or Petange or Peiteng or Petingen or Ettelbruck or Ettelbreck or Ettelbrueck or Dikrech or Strassen or Stroossen or Bertrange or Bartreng or Bartringen or Latgalia or Ogre or "Valle d Aosta" or "Vallee d Aoste Venetia" or "Aosta Valley" or "Vallee d'Aoste" or "Valle d'Aosta" or "na larmhi" or "an Longfoirt" or Muineachan or "Contae Lu" or "Cill Dara" or "Chill Dara" or "Ath Cliath" or "Chill Mhantain" or "Loch Garman" or "Fine Gall" or "Cill Mhantain" or Ceatharlach or "An Mhi" or Mhuineachain or "An Uaimh" or "Dun Dealgan" or "Droichead Atha" or Mumhain or Dunnya or Dinnygal or "Cuige Laighean" or Connachta or Sord or "Cuige Mumhan" or Laighin or Liatroma or Liatroim or "Dhun na nGail" or Sligeach or Shligigh or "Mhaigh Eo" or "Maigh Eoor Ros comain" or Gaillimh or "na Gaillimhe" or "An Clar" or "an Chlair" or "Thiobraid Arann" or Luimneach or | 17150 |
| Luimnigh or "Chill Chainnigh" or "Cill Chainnigh" or Corcaigh or Ciarrai or Chiarrai or "Tiobraid Arann" or Watterford or "Port Lairge" or "An Longfort" or "Contae na Mi" or "An Cabhan" or "An Cabhain" or "Laoise " or "Uibh Fhaili" or "Ionian island" or "Northern Aegean" or Petthalia or "Southern Aegean" or Cyclades or Piraeus or Pireas or "Aegean island" or "Voreio Aigaiou" or "Oros Athos" or "Perifereia Ipeirou" or "North Alfold" or "South Alfold" or "Stara Zagora" or Balgariya or Balgarija or Rusenka or Sofyiska or "Veliko Tarnovoor" or Suomalaiset or Suomen or "Laane Virumaa" or Eestlased or Eestlane or Finn or Finns or Frederiksberg or Storstrom or Bornholm or Ringkjoberg or Frederiksborg or danes or dane or Holomoc or Cestina or Cech or Engomi or Prague or Eglence or Aglandjia or Aglantzia or "Kato Polemidia" or "Kato Polemidhia" or Aradhippou or Iskele or Strovolos or Lakatamia or Lakadamyia or Kyprioi or Goettingen or Bonn or Gottingen or Mitteledeutschland or Rhein or Rhine or Braunschweig or Oldenburg or "Northern Rhine" or "Seine Saint Denis" or "Outre Mer" or Oise or "Noord-Nauw van Kales" or "Rono Arpes" or "Rose Aups" or "Peitau Charantas" or "Poitou-Cherentes" or "Mieidia Pireneus" or "Broiou al Liger" or "Pays de la Loire" or "Mediodia Pirineos" or Lothringen or "Miegiorn Pireneus" or Lottringe or Lemosin or "Frantche Comte" or Normaundie or Aguiene or Auvernhe or Elsass or Auvernha or Aquitania or Bregogne or Akitania or "Lengadoc Rosselhon" or "Llenguadoc-Rossello" or "Franche Comtat" or Lahti).ti.ab. |  |
| 41 (Lunnonderie or Dyfed or Merionethshire or Anthrim or Radnorshire or Montgomeryshire or Aontroim or "Contae Aontroma" or Entrim or "Ard Mhacha" or Airmagh or "An Dun" or "an Duin" or Doon or Doun or "Fear Manach" or "Fhear Manach" or Fermanay or Doire or Dhoire or Banffshire or Berwickshire or Cromartysire or Dumfriesshire or Haddingtonshire or Kincardineshire or Inverness or Dumbarton or Kirkcudbrightshire or Nairnshire or Peeblesshire or Perthshire or Elginshire or Ross* or Roxburghshire or Selkirkshire or Linlithgowshire or Stirlingshire or Zetland or Wigtownshire or Brecknockshire or Caernarfonshire or Clwyd or Cardiganshire or Great Britain or GB or Scots or Cymru or "Isle of Ely" or Cleveland or Cornwall or Hereford or Huntingdon or Humberside or Huntingdonshire or Worcester or Middlesex or Salop or Westmorland or "Isle of Man" or Jersey or Guernsey or "Channel Islands" or Forfarshire or Swedes or Swede or Sodermanlands or "Balear Islands" or Arago or Euskadi or "Baske region" or Galiza or "Las Palmas" or Slovenci or Slovene or Jugovzhodna or Drava or Savinja or Mura or "Central Sava" or Posavska or "Lower Sava" or Laibach or Lubiana or Creina or Carnium or "Marburg an der Drau" or Chreina or Krainbur or Kopar or Wollan or Woellan or Neustadt or Slovaci or Slovenki or Posonium or Besztercebanya or Neutra or Nyitra or Nyitria or Kaschau or Kassa or Presporok or Pressburg or Neusohl or Nagyszombat or Tyrnau or Trencschin or Trencsen or Zvolen or Deutschendorf or Turocszentmarton or Sillein or Tyrnavia or Varat or Temesva or Timisvar or Varad or Jassy or Lassay Galatz or Galac or Tomis or Konstantia or Kostence or Klausenburg or Kolozsvár or Kronstadt or Brasso or Brassovia or Coron or Rumani or Romani or "Alto Tras-os-Montes" or Oporto or Cimbra or Maltin or "s-Gravenhage" or "Den Haag" or Frisia or "Ill Belt" or Birkirkara or "B Kara" or Birchircara or Mosta or Qormi or "St Paul s Bay" or "St Paul's Bay" or "Pawl il Bahar" or Zabbar or Sliema or Naxxar or Gwann or "St John" or Zebbug or "Citta rohan" or Fgura).ab.ti. | 59028 |
| 42 or/32-41 | 3026218 |
| 43 15 and 22 and 27 and 31 and 42 | 385 |
| 44 limit 43 to yr="2020-Current" | 378 |
| 45 15 and 22 and 27 and 31 | 1260 |
| 46 limit 45 to yr="2020-Current" | 1180 |
| Database: Embase <1974 to 2022 January 19> |  |
| **KEY TERMS** | **HITS** |
| 1 exp Coronavirus/ | 8442 |
| 2 Coronaviridae Infections/ or Coronaviridae/ or SARS-CoV-2/ or COVID-19/ | 75117 |

|  |  |  |
| --- | --- | --- |
| 3 | Betacoronavirus 1/ or Betacoronavirus/ | 7527 |
| 4 | Coronavirus Infections/ | 11415 |
| 5 | SARS Virus/ | 7261 |
| 6 | Severe Acute Respiratory Syndrome/ | 10368 |
| 7 | ((severe adj acute adj respiratory adj syndrome) or SARs or Sars-cov or ((sars-associated or sars-related) adj (cov or coronavirus))).mp. | 111209 |
| 8 | SARS-CoV-2/ | 21330 |
| 9 | exp Coronavirus/ or exp Coronavirus Infections/ | 220062 |
| 10 | (Coronavir* or nCov or covid or covid-19).ti,ab,kf. | 221653 |
| 11 | Coronavirus OC43, Human/ | 590 |
| 12 | HKU1.mp. | 713 |
| 13 | HCV-OC43.mp. | 34 |
| 14 | ((("2019" adj (novel or new) adj corona*) or ("2019" adj (CoV or nCoV)) or (coronavirus adj (disease adj "2019")) or COVID19 or COVID-19 or ((Novel or New) adj Corona*) or SARS2 or SARS-CoV-2 or (SARS adj2 (coronaviridae or coronavirus)) or ((sars or Coronavirus) adj "2") or nCov or 2019ncov).mp. | 238025 |
| 15 | or/1-14 | 264506 |
| 16 | (attack rate* or (secondary adj2 attack rate*) or (contact adj2 attack rate*)).mp. | 5647 |
| 17 | Contact Tracing/ or contact*.mp. or (contact* adj1 transmission*).mp. | 558932 |
| 18 | (cluster* or close*adj1 contact*).mp. | 520203 |
| 19 | (second* adj transmission*).mp. | 712 |
| 20 | (contact adj transmission).mp. | 569 |
| 21 | ((transmit* adj1 rate*) or (transmiss* adj1 rate*)).mp. | 6246 |
| 22 | or/16-21 | 1075687 |
| 23 | family.mp. | 1278196 |
| 24 | relatives.mp. | 82841 |
| 25 | house*.mp. | 286819 |
| 26 | home.mp. | 427356 |
| 27 | or/23-26 | 1936966 |
| 28 | young*.mp. | 1363140 |
| 29 | (toddler* or preschool* or child* or pediat* or paediat* or kid or kids or prepubescen* or prepuberty* or puberty or pubescen* or teen* or young* or youth* or minors* or under ag* or underag* or juvenile* or girl* or boy* or preadolesc* or adolesc* or nursery or prekindergarten or kindergarten* or early childhood education or preschool* or elementary education or elementary school* or primary education or primary school* or K-12* or K12 or 1st-grade* or first-grade* or grade 1 or grade one or 2nd-grade* or second-grade* or grade 2 or grade two or 3rd-grade* or third-grade* or grade 3 or grade three or 4th-grade* or fourth-grade* or grade 4 or grade four or 5th-grade* or fifth-grade* or grade 5 or grade five or 6th-grade* or sixth-grade* or grade 6 or grade six or intermediate general or middle school* or secondary education or secondary school*OR 7th-grade* or seventh-grade* or grade 7 or grade seven or 8th-grade* or eight-grade* or grade 8 or grade eight or 9th-grade* or ninth-grade* or grade 9 or grade nine or 10th-grade* or tenth-grade* or grade 10 or grade ten or 11th-grade* or eleventh-grade* or grade 11 or grade eleven or 12th-grade* or twelfth-grade* or grade 12 or grade twelve or junior high* or highschool* or high school* or preuniversity or pre-university or college* or tertiary education or tertiary school*OR postsecondary education or postsecondary school* or prevocational or vocational or classroom* or curricul* or education* or learner* or lesson* or pupi* or school* or student*).ti,ab,kf. | 4728511 |
| 30 | toddlers/ or preschool children/ or young children/ or children/ or pediatrics/ or preadolescents/ or youth/ or | 2060905 |
| adolescents/ or early adolescents/ or late adolescents/ or nursery schools/ or kindergarten/ or early childhood education/ or preschool education/ or preschool teachers/ or elementary secondary education/ or grade 1/ or grade 2/ or grade 3/ or grade 4/ or grade 5/ or grade 6/ or grade 7/ or grade 8/ or grade 9/ or grade 10/ or grade 11/ or grade 12/ or elementary education/ or elementary schools/ or elementary school students/ or elementary school teachers/ or primary education/ or public schools/ or public school teachers/ or middle schools/ or middle school students/ or junior high schools/ or junior high school students/ or secondary education/ or secondary schools/ or secondary school students/ or secondary school teachers/ or high schools/ or high school students/ or college students/ or colleges/ or two year college students/ or two year college students/ or vocational education/ or vocational schools/ or students/ |  |
| 31 or/28-29 | 5012613 |
| 32 exp Europe/ | 1673844 |
| 33 European Union/ | 29014 |
| 34 (europa or europe* or EU or EEA or Euratom or Eurozone or EEC or ECSC or Euroregion or (Schengen and (area or countr* or region* or state*))) .ti,ab. | 576357 |
| 35 (balkan* or baltic* or (mediterranean and (area or countr* or region* or state*)) or (alpine and (area* or countr* or region* or state*)) or nordic* or scandinavia* or danubian or "iberian peninsula" or "peninsula iberi*" or iberica or "iberiar peninsula" or yugoslavia or jugoslavija or jugoslavija or jugoslavia or Ceskoslovensko or "Cesko slovensko" or benelux or fennoscandia or fennoskandi or visegrad* or "grupa wyszehradzka" or "vysehradzka skupina" or "vysehradzka stvorkaor" or "united kingdom" or uk or britain or british or (england not new england) or english or scotland or scottish or wales or welsh or "northern ireland" or london or "east midlands" or "west midlands" or yorkshire or "east anglia" or bedfordshire or hertfordshire or essex or peterborough or cambridgeshire or norfolk or suffolk or luton or bedford or "southend on sea" or thurrock or derbyshire or nottinghamshire or leicestershire or rutland or lincolnshire or derby or leicester or northamptonshire or nottingham or "tyne and wear" or "tees valley" or durham or darlington or hartlepool or "stockton on tees" or northumberland or teesside or sunderland or cumbria or cheshire or manchester or lancashire or merseyside or blackburn or darwen or blackpool or chester or liverpool or sefton or warrington or wirral or berkshire or buckinghamshire or oxfordshire or hampshire or "isle of wight" or kent or surrey or sussex or brighton or hove or "milton keynes" or portsmouth or southampton or devon or dorset or somerset or gloucestershire or wiltshire or bath or bournemouth or poole or bristol or plymouth or swindon or torbay or herefordshire or staffordshire or birmingham or coventry or dudley or sandwell or shropshire or solihull or "stoke on trent" or telford or wrekin or walsall or warwickshire or wolverhampton or worcestershire or barnsley or doncaster or rotherham or bradford or calderdale or kirklees or kingston or leeds or sheffield or wakefield or (york not new york) or antrim or ards or armagh or ballymena or ballymoney or banbridge or carrickfergus or castlereagh or coleraine or cookstown or craigavon or (down and (district or council)) or dungannon or fermanagh or larne or limavady or lisburn or magherafelt or moyle or "newry and mourne" or newtownabbey or omagh or strabane or londonderry or tyrone or belfast or aberdeen or aberdeenshire or angus or dundee or argyll or bute or clackmannanshire or fife or ayrshire or dunbartonshire or lothian or renfrewshire or edinburgh or falkirk or glasgow or highland* or inverclyde or midlothian or moray or lanarkshire or kinross* or stirling or "orkney islands" or "eileanan siaror shetland islands" or bridgend or "neath port talbot" or cardiff or "vale and glamorgan" or "central valleys" or conwy or denbighshire or flintshire or wrexham or "gwent valleys" or gwynedd or "isle and anglesey" or monmouthshire or newport or powys or swansea or ceredigion or carmarthenshire or pembrokeshire or "merthyr tydfil" or "rhondda cynon taff" or "blaenau gwent" or caerphilly or torfaen or caithness or sutherland or cromarty or teeside or tyneside or wearside or "west mercia" or avon or ulster or derry or midway or "east riding" or "west riding" or "lake district" or "peak district" or cumberland or dartmoor or exmoor or sweden or sverige or swedish or svenska or stockholm* or norrland or svealand or mellansverige or smaland or sydsverige or vastsverige or orebro or ostergotland* or | 1156630 |
| vastergotland* or skara* or bohus* or dalsland or narke or sodermanland or uppsala or uppland or vastmanland* or jamtland* or harjedalen or vasternorrland* or dalarna or kopparberg or gavleborg* or gastrikland or halsingland or varmland* or gotland* or oland or jonkoping* or kalmar* or kronoberg* or blekinge or skane* or norrbottn* or vasterbotten* or lappland or angermanland or medelpad or halland* or gotaland* or gothenburg or goteborg* or malmo* or vasteras or linkoping or helsingborg or halsingborg or norrkoping or gavle or umea or lulea or karlstad or kalmar or huddinge or solna or ostersjo* or malaren* or malardalen or spain or espana or spanish or espanol* or spaniard* or madrid or andalucia or andalusia).ab,ti. |  |
| 36 (aragon or cantabria or canarias or "canary islands" or "castilla y leon" or "castile la mancha" or "castilla la mancha" or cataluna or catalonia or ceuta or melilla or navarra or navarre or valencia* or extremadura or galicia or balears or "balearic islands" or baleares or "la rioja" or "pais vasco" or "basque country" or coruna or alava or araba or albacete or alicante or alacant or almeria or asturias or avila or badajoz or badajos or barcelona or burgos or caceres or cadiz or castellon or castello or "ciudad real" or (cordoba not argent*) or cuenca or eivissa or ibiza or formentera or "el hierro" or fuerteventura or girona or gerona or "gran canaria" or granada or (guadalajara not mexic*) or guipuzcoa or gipuzkoa or huelva or huesca or jaen or gomera or palma or lanzarote or leon or lleida or lerida or lugo or malaga or mallorca or majorca or menorca or minorca or murcia or ourense or orense or palencia or pontevedra or salamanca or segovia or sevilla or seville or soria or tarragona or tenerife or teruel or toledo or valencia or valladolid or vizcaya or biscay or zamora or zaragoza or saragossa or bilbao or bilbo or compostela or "san sebastian" or donostia or vitoria or oviedo or pamplona or logrono or gasteiz or slovenia* or slovenija or ljubljana or gorenjska or carniola or goriska or gorizia or koroska or carinthia or "notranjsko kraska" or "obalno kraska" or "coastal karst" or podravska or pomurska or savinjska or spodnjeposavska or zasavska or osrednjeslovenska or maribor or celje or kranj or velenje or koper or capodistria or "novo mesto" or ptuj or trbovlje or kamnik or murska or sobota or "nova gorica" or slovakia or slovensk* or slovak* or bratislav* or nitria* or nitra or trencian* or trencin or banskobystri* or "banska bystrica" or zilina or zilin* or trnava or trnav* or presov* or kosic* or (martin and (city or svaty)) or poprad or italy or italia* or rome or roma or abruzzo or abruzzu or basilicata or lucania or calabria or campania or "emilia romagna" or "friuli venezia giulia" or lazio or latium or liguria* or lombardy or lombardia or marche or marches or molisano or molise or piedmont* or piemonte or sardinia or sardegna or sicily or sicilia or toscana or tuscanly or trentino or trento or umbria or veneto or triveneto or puglia or apulia or bolzano or bozen or milan or milano or naples or napoli or turin or torino or palermo or genoa or genova or florence or firenze or bari or catania or venezia or venice or padova or padua or siena or bologna or trieste or urbino or aosta or aoste or perugia or brescia or cagliari or catanzaro or aquila or ancona or ireland or ireland or eire or irish* or dublin or fingsal or "dun laoghaire" or wicklow or wexford or carlow or kildare or meath or louth or monaghan or cavan or longford or westmeath or offaly or laois or kilkenny or waterford or cork or kerry or limerick or tipperary or clare or galway or mayo or roscommon or sligo or leitrim or donegal or drogheda or dundalk or swords or bray or navan or leinster or connacht or hungar* or budapest or transdanubia or magyarország or magyar or dunantuli or dunantul or "great plain" or "alföld es eszak" or "eszak alföld" or "del alföld" or bacs or kiskun or "northern alföld" or "southern alföld" or baranya or bekes or borsod or abauj or zemplen or foharos or csongrad or fejer or moson or sopron or hajdu or bihar or heves or "jász nagykn szolnok" or komarom or esztergom or nograd or (Pest and (megye or county)) or somogy or szabolcs or szatmar or bereg or tolna or vas or veszprem or zala or zalaegerszeg or debrecen or miskolc or szeged or pecs or gyor or nyiregyhaza or kecskemet or szekesfehervar or szombathely or bekescsaba or eger or tatanya or salgotarjan or kaposvar or szekszard or greece or "hellenic republic" or greek* or ellada or "elliniki dimokratia" or hellas or hellenes or attica or attiki or makedonia or macedonia or thraki or thrace or crete or kriti or epirus or ipeiros or "ionia nisia" or "ionian neson" or "ionian islands" or "north aegean" or "aegean islands" or "nisi aiagiu" or "notio aigaiu" or peloponnese or peloponnisos or "voreio aigaiu" or "south aegean" or thessaly or thessalia).ab,ti. | 660919 |
| 37 (cycklades or kiklades or dodecanese or dodekanisa or "mount athos" or "omicronros alphathos" or athens or athina or thessaloniki or thessalonica or patras or patra or heraklion or heraclion or iraklion or irakleion or iraklio or larissa or larisa or volos or rhodes or rodos or ioannina or janina or yannena or chania or chalcis or chalkida or alexandroupoli or german* or deutschland or deutsch* or bundesrepublik or westdeutschland or ostdeutschland or baden or wuerttemberg or wurtemberg or bayern or bavaria or berlin or brandenburg or bremen or hamburg or hessen or hesse or hessia or mecklenburg or vorpommern or pomerania or niedersachsen or neddersassen or saxony or niederbayern or "north rhine" or westphalia or westfalen or "rhineland palatinate" or "rheinland pfalz" or saarland or sachsen or "schleswig holstein" or thuringia or thuringen or thueringen or freiburg or karlsruhe or callsruhe or stuttgart or tuingen or oberbayern or "upper palatinate" or oberpfalz or franken or franconia or oberfranken or mittelfranken or schwaben or unterfranken or swabia or darmstadt or giessen or kassel or arnsberg or cologne or koln or koeln or detmold or dusseldorf or duesseldorf or munster or muenster or munich or munchen or muenchen or frankfurt or dortmund or essen or numberg or nuernberg or nuremberg or hanover or hannover or leipzig or dresden or ruhrgebiet or revier or ruhrpott or pott or ruhr or france or french* or francais or alsace or aquitaine or auvergne or brittany or bretagne or bourgogne or burgundy or "champagne ardenne" or "franche comte" or "ile de france" or "languedoc roussillon" or limousin or lorraine or normandie or normandy or "midi pyrenees" or "nord pas de calais" or picardie or picardy or "poitou charentes" or provence or "rhone alpes" or corse or corsica or guiana or guyane or guadeloupe or martinique or reunion or mayotte or ain or aise or allier or "alpes de haute provence" or "haute alpes" or "alpes maritimes" or ardeche or ardennes or ariege or aube or aude or aveyron or "bas rhin" or "bouches du rhone" or calvados or cantal or charente or cher or correze or "corse du sud" or cote* or azur* or creuse or "deux sevres" or dordogne or doubs or drome or essonne or eure or finistere or gard or gers or gironde or "haute corse" or "haute garonne" or "haute marne" or "hautes alpes" or "haute saone" or "haute savoie" or "hautes pyrenees" or "haute vienne" or "haut rhin" or "hauts de seine" or herault or "ille et vilaine" or indre or isere or jura or landes or loire or loiret or (lot and (departement or department)) or "lot et garonne" or "loir et cher" or lozere or manche or marne or mayenne or "meurthe et moselle" or meuse or morbihan or moselle or (nord and (department or departement)) or nievre or oise or orne or "pas de calais" or paris or "puy de dome" or "pyrenees atlantiques" or "pyrenees orientales" or rhone or sarthe or savoie or "seine et marne" or "seine maritime" or somme or tarn or "territoire de belfort" or "val de marne" or var or vaucluse or vendee or vienne or vosges or yonne or yvelines or marseille or lyon or nice or nantes or strasbourg or montpellier or bordeaux or lille or toulouse or finland or finnish* or suomi* or lapland or lappi or lapland or ostrobothnia or pohjanmaa or osterbotten or kainuu or kajanaland* or karelia or karjala or karelen or savonia or savo or savolax or pirkanmaa or birkaland or satakunta or satakunda or tavastia or tavastland or "pajjat hame" or "kanta hame" or uusimaa or nyland or kymenlaakso or kymmenedalen or aland or ahvenanmaa or helsinki or helsingfors or espoo or esbo or tampere or tammerfors or vantaa or vanda or oulu or uleaborg or turku or abo or jyvaskyla or kuopio or lathi or lahtis or kouvola or estonia* or eesti or esti or tallinn or harju or harjumaa or hiiu or hiiumaa or "ida viru" or "ida virumaa" or jarvamaa or jarva or jogevamaa or jogeva or laanemaa or laane or parnumaa or polva or polvamaa or rapla or raplamaa or saare or saaremaa or tartu or tartumaa or valga or valgamaa or valgamaakond or viljandimaa or voru or vorumaa or narva or parnu or kohtla jarve or viljandi).ab.ti. | 708042 |
| 38 (rakvere or maardu or sillamae or kuressaare or romania* or rumania* or roumania* or romanian or roman or bucharest or Bucuresti or alba or brasov or covasna or harghita or mures or sibiu or bacau or botosani or iasi or neamt or suceava or vaslui or bihor or "bistrita nasaud" or cluj or maramures or salaj or "satu mare" or arges or calarasi or dambovita or giurgiu or ialomita or prahova or teleorman or braila or buzau or galati or tulcea or vrancea or dolj or gorj or mehedinti or (olt and (river or county or region or judetul or raul)) or valcea or vilcea or arad or caras-severin or hunedoara or timis or ilfov or timisoara or constanta or craiova or ploiesti or oradea or cluj- napoca or deva or portugal or portugues* or lisboa or lisbon or leira or santarem or beja or faro or evora or | 458552 |
| portalegre or "castelo branco" or guarda or aveiro or viseu or braganca or "vila real" or "viana do castelo" or alentejo or azores or acores or madeira or "os montes" or (ave and (community or intermunicipal or comunidade)) or mondego or vouga or beira or cavado or lafoes or douro or porto or tejo or minho or setubal or pinhal or "serra da estrela" or tamega or algarve or gaia or amadora or braga or (agualva and cacem) or funchal or coimbra or almada or poland or polska or polish or polski or pole or poles or polak or polka or polacy or warsaw or warszawa or wielkopolskie or pomerania* or pomorskie or kuyavian or kujawsko or malopolskie or lodz or lodzkie or silesia* or dolnoslaskie or lublin or lubelskie or lubus or lubusz or lubuskie or masovia or mazowske or masovian or mazowieckie or opole or opolskie or podlaskie or podlachia or podlasie or subcarpathian* or carpathian* or podkarpackie or swietokrzyskie or slaskie or slask or "varmia mazuria" or "varmian mazurian" or "varmia masuria" or "varmian masurian" or "warmia mazury" or "warminsko mazurskie" or zachodniopomorskie or krakow or cracow or wroclaw or poznan or gdansk or szczecin or bydgoszcz or katowice or bialystok or olsztyn or kielce or "zielona gora" or torun or "gorzow wielkopolski" or netherlands or nederland* or dutch* or amsterdam or drenthe or flevoland or friesland or frysian or gelderland or guelders or groningen or limburg or "north brabant" or "noord brabant" or holland or overijssel or overijssel or utrecht or zeeland or rotterdam or hague or eindhoven or tilburg or almere or breda or nijmegen or nimeguen or malta or maltese or valletta or gozo or ghawdex or luxembourg* or luxemburg or letzebuerg or diekirch or grevenmacher or lithuania* or "lietuvos respublika" or lietuva or lietuviu or vilnius or vilniaus or kaunas or kauno or klaipeda or klaipedos or panevezys or panevezio or siauliai or siauliu or alytus or alytaus or taurages or taurage or marijampoles or marijampole or telsiu or telsiai or utenos or utena or mazeikiai or jonava or mazeikiu or jonavos or latvi* or latvija* or riga or courland or kurzeme or kurland or latgale or lettgallia or latgola or vidzeme or vidumo or semigallia or semigalia or zemgale or pieriga or daugavpils or dinaburg or liepaja or libau or jelgava or jurmala or jekabpils or jakobstadt or rezekne or rezne or rositten or valmiera or wolmar or ventspils or windau or denmark or danish* or danmark or dansk* or hovedstaden or midtjylland or sjælland or sealand or syddanmark or jutland or jylland or nordjylland or sonderjyllands or "zealand region" or "region zealand" or hillerod or viborg or aalborg or alborg or soro or vejle or copenhagen or kobenhavn or arhus or aarhus or roskilde or odense or frederiksberg or esbjerg or gentofte or gladsaxe or randers or kolding or czech* or cesk* or stredoces* or jihoce* or bohemia or bohemian or kralovehradec* or "hradec kralove" or karlovars* or "karlovy vary" or liberec* or moravskoslezs* or "moravian silesian" or olomouc* or pardubic* or pardubice or plzen* or pilsen or prague or praha or prag or jihomorav* or moravia or moravian or morava or usteck* or usti or vysocina or zlin or zlinsk* or "ceske budejovice" or budweis or brno or ostrava or cyprus or cypriot* or kypros or kibris or kypriaki* or nicosia or lefkosa).ab,ti. |  |
| 39 (lefkosia or famagusta or magusa or ammochoostos or gazimagusa or kyrenia or girne or keryneia or larnaca or larnaka or limassol or lemesos or limasol or leymosun or paphos or pafos or baf or gazibaf or protaras or pergamos or beyarmudu or morfou or guzelyurt or omorfo or morphou or aradippou or croatia* or hrvatsk* or hrvati or bjelovar or "bjelovarsko bilogorska" or "brod posavina" or "brodsko posavska" or "dubrovnik neretva" or "dubrovačko neretvanska" or istria or istarska or karlovacka or karlovac or "kopriivnicko krizevacka" or kopriivnica or krizevci or "krapina zagorje" or "krapinsko zagorska" or "lika senj" or "licko senjska" or medimurska or medimurje or osijek or osjecko or baranja or "osjecko baranjska" or "pozega slavonia" or "pozesko slavonska" or "primorje gorski kotar" or "primorsko goranska" or "sibensko kninska" or "sibensko kninske" or sibenik or knin or sisak or "sisacko moslavacka" or moslavina or "splitsko dalmatinska" or split or dalmatia or varazdin or varazdinska or viroviticko-podravska or virovitica or podravina or "vukovarsko srijemska" or vukovar or srijem or zadar or zadarska or zagreb or zagrebacka or rijeka or "velika gorica" or "slavonski brod" or pula or bulgaria* or sofia or gabrovo or blagoevgrad or "pirin macedonia" or burgas or dobrich or haskovo or kardzhali or kurdzhali or kyustendil or lovech or montana or pazardzhik or pernik or pleven or plovdiv or razgrad or rousse or ruse or shumen or sliven or silistra or smolyan or "stara zagora" or targovishte or varna or tarnovo or vidin or vratsa or vratza or yambol or belgi* or belge or belgisch or brussel* or bruxelles or bruxelloise or flemish or flamand or | 581171 |
| flemisch or flanders or flandern or flandre or vlaanderen or vlaams or flamande or waals or walloon* or wallon* or antwerp* or anvers or ostflandern or "vlaams brabant" or limburg or limburg or hainault or hainaut or henegouwen or hennegau or liege or luik or luttich or namur or namen or westflandern or "waals brabant" or ghent or gent or gand or charleroi or bruges or brugge or schaerbeek or schaarbeek or anderlecht or leuven or louvain or hasselt or mons or wavre or waver or austria* or vienna or wien or osterreich* or sudosterreich or westosterreich or niederosterreich or burgenland or carinthia or karnten or oberosterreich or styria or steiermark or salzburg or saizburg or tyrol or tirol or becs or vorarlberg or bregenz or linz or eisenstadt or innsbruck or graz or klagenfurt or polten or villach or wels or dornbirn or feldkirch or steyr or iceland or icelandic* or islenska* or icelander* or islendinga* or islendigar or inslenska or reykjavik or reykjavikurborg or hofudborgarsvaedid or sudurnes or vesturland or vestfirðir or westfjords or nordurland or austurland or sudurland or kopavogur or hafnarfjörður or Akureyri or Gardabaer or Mosfellsbaer or Keflavik or Akranes or Selfoss or Seltjarnarnes or switzerland or schweiz or schweizerische or swiss or suisse or aargau or argovia or ausserrhoden or "outer rhodes" or innerrhoden or "inner rhodes" or basel or bern or berne or fribourg or freiburg or geneva or geneve or glarus or graubunden or grisons or grigioni or jura or lucerne or luzern or neuchatel or zurich or (uri and (canton or kanton)) or schwyz or obwalden or nidwalden or zug or solothurn or schaffhausen or thurgau or thurgovia or ticino or tessin or vaud or valais or wallis or "st gallen" or lausanne or winterthur or winterthour or lugano or biel or bienne or norway or norwegian* or norge or noreg or norgga or ostfold or akershus or oslo or hedmark or oppland or buskerud or vestfold or telemark or "aust agder" or "vest agder" or rogaland or hordaland or "sogn og fjordane" or "sogn and fjordane" or "sogn fjordane" or "more og romsdal" or "more and romsdal" or "more romsdal" or trondelag or nordland or troms or finnmark or bergen or stavanger or sandnes or trondheim or kristiansand or drammen or fredrikstad or sarpsborg or porsgrunn or skien or tonsberg or alesund or liechtenstein or vaduz or triesenberg or triesen or schellenberg or schaan or ruggell or planken or mauren or gamprin or eschen or balzers or turkey or turkiye or turkish or istanbul or marmara or aegean or anatolia or "black sea" or tekirdag or balikeseir or izmir or aydin or manisa or bursa or kocaeli or ankara or konya or antalya or adana or hatay or kirikkale or kayseri or zonguldak or kastamonu or samsun or trabzon or erzurum or agri or malatya or (van and (region or subregion or bolgesi)) or gaziantep or sanliurfa or mardin or mersin).ti.ab. |  |
| 40 ("Esch sur Alzette" or "Esch Uelzecht" or "Esch an der Alzette" or "Esch an der Alzig" or Dudelange or Diddeleng or Dudelingen or Dueelingen or Schiffange or Scheffleng or Schifflingen or Bettembourg or Beetebuerg or Bettemburg or Petange or Peiteng or Petingen or Ettelbruck or Ettelbreck or Ettelbrueck or Dikrech or Strassen or Stroossen or Bertrange or Bartreng or Bartringen or Latgalia or Ogre or "Valle d Aosta" or "Vallee d Aoste Venetia" or "Aosta Valley" or "Vallee d'Aoste" or "Valle d'Aosta" or "na larmhi" or "an Longfoirt" or Muineachan or "Contae Lu" or "Cill Dara" or "Chill Dara" or "Ath Cliath" or "Chill Mhantain" or "Loch Garman" or "Fine Gall" or "Cill Mhantain" or Ceatharlach or "An Mhi" or Mhuineachain or "An Uaimh" or "Dun Dealgan" or "Droichead Atha" or Mumhain or Dunnya or Dinnygal or "Cuige Laighean" or Connachta or Sord or "Cuige Mumhan" or Laighin or Liatroma or Liatroim or "Dhun na nGall" or Sligeach or Shligigh or "Mhaigh Eo" or "Maigh Eoor Ros comain" or Gaillimh or "na Gaillimhe" or "An Clar" or "an Chlair" or "Thiobraid Arann" or Luimneach or Luimnigh or "Chill Chainnigh" or "Cill Chainnigh" or Corcaigh or Ciarrai or Chiarrai or "Tiobraid Arann" or Watterford or "Port Lairge" or "An Longfort" or "Contae na Mi" or "An Cabhan" or "An Cabhain" or "Laoise " or "Uibh Fhaili" or "Ionian island" or "Northern Aegean" or Petthalia or "Southern Aegean" or Cyclades or Piraeus or Pireas or "Aegean island" or "Voreio Aigaiou" or "Oros Athos" or "Perifereia Ipeirou" or "North Alfold" or "South Alfold" or "Stara Zagora" or Balgariya or Balgarija or Rusenka or Sofyiska or "Veliko Tarnovoor" or Suomalaiset or Suomen or "Laane Virumaa" or Eestlased or Eestlane or Finn or Finns or Frederiksberg or Storstrom or Bornholm or Ringkjobing or Frederiksborg or danes or dane or Holomoc or Cestina or Cech or Engomi or Prague or Eglence or Aglandjia or Aglantzia or "Kato Polemidia" or "Kato Polemidhia" or Aradhippou or Iskele or Strovolos or Lakatamia or Lakadamya or Kyprioi or Goettingen or Bonn or Gottingen or Mitteldeutschland or | 23298 |
| Rhein or Rhine or Braunschweig or Oldenburg or "Northern Rhine" or "Seine Saint Denis" or "Outre Mer" or Oise or "Noord-Nauw van Kales" or "Rono Arpes" or "Rose Aups" or "Peitau Charantas" or "Poitou-Cherentes" or "Miedia Pireneus" or "Broiou al Liger" or "Pays de la Loire" or "Mediodia Pirineos" or Lothringen or "Miegjorn Pireneus" or Lottringe or Lemosin or "Frantche Comte" or Normaundie or Aguiene or Auvernhe or Elsass or Auvernha or Aquitania or Bregogne or Akitania or "Lengadoc Rosselhon" or "Llenguadoc-Rossello" or "Franche Comtat" or Lahti).ti.ab. |  |
| 41 (Lunnonderrie or Dyfed or Merionethshire or Anthrim or Radnorshire or Montgomeryshire or Aontroim or "Contae Aontroma" or Entrim or "Ard Mhacha" or Airmagh or "An Dun" or "an Duin" or Doon or Doun or "Fear Manach" or "Fhear Manach" or Fermanay or Doire or Dhoire or Banffshire or Berwickshire or Cromartyshe or Dumfriesshire or Haddingtonshire or Kincardineshire or Inverness or Dumbarton or Kirkcudbrightshire or Nairnshire or Peeblesshire or Perthshire or Elginshire or Ross* or Roxburghshire or Selkirkshire or Linlithgowshire or Stirlingshire or Zetland or Wigtownshire or Brecknockshire or Caernarfonshire or Clwyd or Cardiganshire or Great Britain or GB or Scots or Cymru or "Isle of Ely" or Cleveland or Cornwall or Hereford or Huntingdon or Humberside or Huntingdonshire or Worcester or Middlesex or Salop or Westmorland or "Isle of Man" or Jersey or Guernsey or "Channel Islands" or Forfarshire or Swedes or Swede or Sodermanlands or "Balear Islands" or Arago or Euskadi or "Baske region" or Galiza or "Las Palmas" or Slovenci or Slovene or Jugovzhodna or Drava or Savinja or Mura or "Central Sava" or Posavska or "Lower Sava" or Laibach or Lubiana or Creina or Carnium or "Marburg an der Drau" or Chreina or Krainbur or Kopar or Wollan or Woellan or Neustadt or Slovaci or Slovenki or Posonium or Besztercebanya or Neutra or Nyitra or Nyitra or Kaschau or Kassa or Presporok or Pressburg or Neusohl or Nagyszombat or Tyrnau or Trencsén or Trencsen or Zvolen or Deutschendorf or Turocszentmarton or Sillein or Tynavia or Varat or Temesva or Timisvar or Varad or Jassy or Lassay Galatz or Galac or Tomis or Konstantia or Kostence or Klausenburg or Kolozsvár or Kronstadt or Brasso or Brassovia or Coron or Rumani or Romani or "Alto Tras-os-Montes" or Oporto or Cimbra or Maltin or "s-Gravenhage" or "Den Haag" or Frisia or "Ill Belt" or Birkirkara or "B Kara" or Birchircara or Mosta or Qormi or "St Paul's Bay" or "St Paul's Bay" or "Pawl il Bahar" or Zabbar or Sliema or Naxxar or Gwann or "St John" or Zebbug or "Citta rohan" or Fgura).ab.ti. | 77622 |
| 42 or/32-41 | 4204562 |
| 43 15 and 22 and 27 and 31 and 42 | 467 |
| 44 limit 43 to yr="2020-Current" | 454 |
| 45 15 and 22 and 27 and 31 | 1539 |
| 46 limit 45 to yr="2020-Current" | 1451 |

**Supplementary Table 2.** Descriptive characteristics of included studies (n=26)

| *Author* | *Country, area* | *Study design* | *Timeframe* | *COVID-19 diagnosis* | *SARS-CoV-2 strain* | *Parallel NPIs* |
| --- | --- | --- | --- | --- | --- | --- |
| Abbas 2021 (29) | Sweden | Case study | 4 April 2020 - 8 May 2020 | PCR and/or serology | Not mentioned | Not mentioned |
| Bistaraki 2021 (9) | Greece | Cohort | From 1 October to 9 December 2020 | PCR | Not mentioned | (i) Mandatory mask use, (ii) Lockdown on 7 November 2020, (iii) Cases were instructed to isolate for 10 days |
| Calvani 2021 (32) | Italy | Case-control | October 16 to December 19, 2020 | Ag RDT, NAAT | Not mentioned | Nursery, primary, and middle/junior high schools were opened during our study period, while high schools were opened only until October 26th, when remote teaching was activated. |
| Charbonnier 2021 (10) | Paris, France | Cohort | Between May 8 and July 27, 2020 | RT-PCR and serological RDT | Not mentioned | Positive cases were isolated for 14 days and those with negative test were isolated for 7 days. Protective measures to avoid viral transmission were explained, along with distribution of masks and hydroalcoholic lotions |
| Chudasama 2021 (33) | England | Cross-sectional | August to October 2021 | PCR, LFD | Not mentioned | Easing of COVID-19 restrictions in England in the summer of 2021, including the removal of face coverings or masks from 19 July 2021 and the requirement for self-isolation of close contacts of cases who are children or fully vaccinated. |
| Katlama 2022 (26) | Paris, France | Cross-sectional | July -September 2020 | PCR and/or serology | Not mentioned | Recommended masks |
| Dupraz 2021 (25) | Canton of Vaud, Switzerland | Cross-sectional | February 27 - April 1, 2020 | NAAT | Not mentioned | Not mentioned |
| Galow 2021 (11) | Dresden, Germany | Cohort | June 2020 | PCR | Not mentioned | Temporal separation in the use of common rooms was implemented most commonly followed by mask wearing of the index person |
| Hall 2021 (12) | England | Cohort | April to September 2020 | PCR | Not mentioned | Not mentioned |
| Hare 2021 (30) | Ireland | Case study | 25 June - 15 November 2020 (case study: 13 August-25 August 2020) | PCR | Not mentioned | Not mentioned |
| Julin 2021 (13) | Oslo/Viken, Norway | Cohort | From May to June 2020, and from September 2020 to the end of April 2021 | PCR | Alpha/ Beta / non-VOC viruses | Some wore masks, kept a distance and/or isolated |
| Koureas 2021 (27) | Larissa, Greece | Cross-sectional | 8 April–4 June 2020 | PCR | Not mentioned | (i) large-scale population screening, (iv) contact tracing of confirmed cases, (v) repeated PCR testing of street vendors, (vi) population screening in all major Roma settlements of the region, (vii) movement restrictions and gathering prohibition, (viii) the isolation of confirmed cases in a specified isolation facility |
| Kuwelker 2021 (31) | Bergen, Norway | Case series | From 28th February to 4th April 2020 | PCR | Not mentioned | Index patients were home isolated, and their household members were instructed to quarantine |
| Ladhani 2021 (14) | England | Cohort | May 2020 | PCR and/or serology | Not mentioned | Self-isolation of index cases |
| Loenenbach 2021 (15) | Hesse, Germany | Cohort | January-February 2021 | PCR | SARS-CoV-2 variant B.1.1.7 | Not mentioned |
| Lyngse 2021 (16) | Denmark | Cohort | January 11 to February 7, 2021 | PCR | SARS-CoV-2 variant B.1.1.7 | Not mentioned |
| Maltezou 2020 (17) | Greece | Cohort | From February 26 to June 30, 2020 | PCR | Not mentioned | Contacts' isolation for 14 days |
| Maltezou 2021 (18) | Athens and Thessaloniki, Greece | Cohort | From 26 February to 3 May 2020 | PCR | Not mentioned | Close contacts were isolated for 14 days following the last contact with the COVID-19 case |
| Miller 2021 (19) | England | Cohort | Between 30th March and 17th November 2020 | PCR | Not mentioned | Isolation of cases and quarantine of contacts |
| Nunziata 2021 (20) | Italy | Cohort | April 2020 to February 2021 | PCR | Not mentioned | The caregiver wore a surgical mask and ensured hygiene. Some children wore masks, but estimated use was 50% of the total time. |
| Posfay-Barbe 2020 (21) | Switzerland | Cohort | 10 March to 10 April 2020 | PCR | Not mentioned | Not mentioned |
| Reukers 2021 (22) | The Netherlands | Cohort | March to May 2020 | PCR and/or serology | Not mentioned | Not mentioned |
| Soriano-Arandes 2021 (23) | Catalonia, Spain | Cohort | 1 July to 31 October 2020 | PCR/rapid antigen test | Not mentioned | Children >6 years old wore masks in school |
| Stich 2021 (28) | Germany | Cross-sectional | 11 May to 1 August 2020 | PCR | Not mentioned | Not mentioned |
| Telle 2021 (24) | Norway | Cohort | 1 March 2020 to 1 January 2021 | PCR | Not mentioned | Not mentioned |

**Supplementary Table 3:** Results from the JBI critical appraisal tool for cohort studies (n=16)

| Studies | Q1 | Q2 | Q3 | Q4 | Q5 | Q6 | Q7 | Q8 | Q9 | Q10 | Q11 |
| --- | --- | --- | --- | --- | --- | --- | --- | --- | --- | --- | --- |
| Bistaraki 2021 | YES | YES | YES | YES | YES | YES | YES | UNCLEAR | UNCLEAR | UNCLEAR | YES |
| Charbonnier 2021 | YES | YES | YES | NO | NO | YES | YES | UNCLEAR | UNCLEAR | UNCLEAR | NO |
| Galow 2021 | YES | YES | YES | NO | NO | YES | YES | UNCLEAR | UNCLEAR | UNCLEAR | YES |
| Hall 2021 | YES | YES | YES | YES | YES | YES | YES | YES | YES | UNCLEAR | YES |
| Julin 2021 | YES | YES | YES | YES | YES | YES | YES | YES | YES | NO | YES |
| Ladhani 2021 | YES | YES | YES | NO | NO | YES | YES | UNCLEAR | UNCLEAR | UNCLEAR | YES |
| Loenenbach 2021 | YES | YES | YES | NO | NO | YES | YES | UNCLEAR | UNCLEAR | UNCLEAR | NO |
| Lyngse 2021 | YES | YES | YES | YES | YES | YES | YES | YES | YES | UNCLEAR | YES |
| Maltezou 2020 | YES | YES | YES | NO | NO | YES | YES | UNCLEAR | UNCLEAR | UNCLEAR | YES |
| Maltezou 2021 | YES | YES | YES | NO | NO | YES | YES | UNCLEAR | UNCLEAR | UNCLEAR | NO |
| Miller 2021 | YES | YES | YES | YES | YES | YES | YES | YES | YES | NO | YES |
| Nunziata 2021 | YES | YES | YES | NO | NO | YES | YES | UNCLEAR | UNCLEAR | UNCLEAR | NO |
| Posfay-Barbe 2020 | YES | YES | YES | NO | NO | YES | YES | YES | YES | UNCLEAR | NO |
| Reukers 2021 | YES | YES | YES | YES | YES | YES | YES | YES | YES | UNCLEAR | YES |
| Soriano-Arandes 2021 | YES | YES | YES | YES | YES | YES | YES | YES | YES | UNCLEAR | YES |
| Telle 2021 | YES | YES | YES | NO | NO | YES | YES | YES | YES | UNCLEAR | NO |

**Supplementary Table 4:** Results from the JBI critical appraisal tool for cross-sectional studies (n=6)

| Studies | Q1 | Q2 | Q3 | Q4 | Q5 | Q6 | Q7 | Q8 |
| --- | --- | --- | --- | --- | --- | --- | --- | --- |
| Chudasama 2021 | YES | YES | YES | YES | YES | NO | YES | YES |
| Dupraz 2021 | YES | YES | YES | YES | YES | YES | YES | YES |
| Katlama 2022 | YES | YES | YES | YES | YES | YES | YES | YES |
| Koureas 2021 | YES | YES | YES | YES | YES | YES | YES | YES |
| Stich 2021 | YES | YES | YES | YES | YES | YES | YES | YES |

**Supplementary Table 5:**
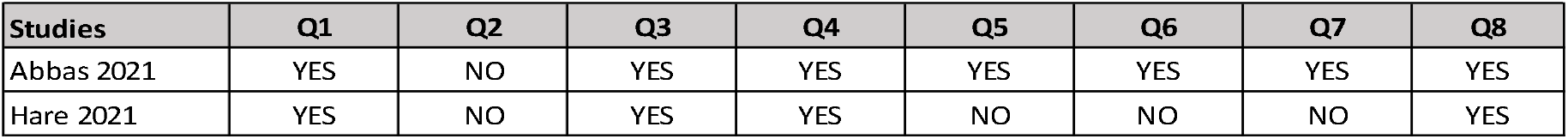
Results from the JBI critical appraisal tool for case reports (n=3)

**Supplementary Table 6:** Results from the JBI critical appraisal tool for case series (n=1)

| Studies | Q1 | Q2 | Q3 | Q4 | Q5 | Q6 | Q7 | Q8 | Q9 | Q10 |
| --- | --- | --- | --- | --- | --- | --- | --- | --- | --- | --- |
| Kuvelker 2021 | YES | YES | YES | YES | YES | YES | YES | YES | YES | YES |

**Supplementary Table 7:** Results from the JBI critical appraisal tool for case series (n=1)

| Studies | Q1 | Q2 | Q3 | Q4 | Q5 | Q6 | Q7 | Q8 | Q9 | Q10 |
| --- | --- | --- | --- | --- | --- | --- | --- | --- | --- | --- |
| Calvani 2021 | YES | YES | YES | YES | YES | YES | YES | YES | YES | YES |

## Notes

### Competing Interest Statement

The authors have declared no competing interest.

